# QUANTIFYING EXPLAINABLE AI METHODS IN MEDICAL DIAGNOSIS: A STUDY IN SKIN CANCER

**DOI:** 10.1101/2024.12.08.24318158

**Authors:** Hardik Sangwan

## Abstract

Deep learning models have shown substantial promise in assisting medical diagnosis, offering the potential to improve patient outcomes and reduce clinician workloads. However, the widespread adoption of these models in clinical practice has been hindered by concerns surrounding their trustworthiness, transparency, and interpretability. Addressing these challenges requires not only the development of explainable AI (xAI) techniques but also quantitative metrics to evaluate their effectiveness.

This study presents a comprehensive framework for training, explaining, and quantitatively assessing deep learning models for skin cancer diagnosis. Leveraging the HAM10000 dataset of seven diagnostic skin lesion categories, multiple convolutional neural network architectures—including custom CNNs, DenseNet, MobileNet, and ResNet—were trained and optimized using augmentation, oversampling, and hyperparameter tuning. Following model training, explainability techniques such as SHAP, LIME, and Integrated Gradients were deployed to generate post hoc explanations. Critically, the primary contribution of this work is the quantitative evaluation of these explanation methods using metrics related to faithfulness, robustness, and complexity. All code, models, and results are publicly available, providing a reproducible pathway toward more trustworthy, explainable diagnostic tools.

## 1 Introduction

Recent advances in machine learning and deep learning have enabled the development of highly accurate diagnostic models in various medical domains, including neurology, oncology, dermatology, and gait analysis. While these models have produced impressive results, their adoption in real-world clinical settings remains limited. Key concerns include the models’ generalization abilities and the limited trust clinicians place in tools whose decision-making processes are often opaque. As healthcare increasingly relies on data-driven support tools, enhancing the interpretability and explainability of these models has become an urgent priority. Explainable AI (xAI) techniques have emerged as a critical avenue for building trust, promoting transparency, and ensuring that healthcare professionals can understand the reasoning behind a model’s predictions. However, a critical gap exists in the quantitative assessment of these techniques. Although numerous studies have applied xAI methods—such as feature importance visualizations—few have systematically measured their effectiveness. Without such quantitative benchmarks, it remains challenging to refine xAI methods, compare their utility, and identify the best approaches for specific clinical tasks.

Let’s look at dermatological applications for instance. Skin lesions, including malignant melanomas, pose a significant global health concern. Early detection is crucial: while localized melanomas have a 98% five-year survival rate, metastatic cases can drop as low as 18%. Dermatological expertise is required to assess suspicious lesions, a process that can be labor-intensive and prone to human variability. Automated systems that can accurately identify and classify lesions offer the promise of earlier, more efficient diagnoses, potentially reducing the need for invasive and time-consuming biopsy procedures. Leveraging the HAM10000 dataset, researchers have achieved impressive classification performance for various lesion types using both traditional machine learning and advanced deep learning models (e.g., DenseNet, MobileNet, ResNet) (Tschandl et al., 2018). Many of these methods are nearing translational feasibility, particularly as a growing number of AI-based tools are being approved by standards agencies and as medical curricula begin to incorporate machine learning concepts.

However, despite models achieving accuracies that match or even surpass human experts (Haenssle et al., 2018; Brinker et al., 2019), many concerns persist about the reliability, interpretability, and transparency of these models.

A medical expert cannot easily reason through the output of a “black box” model. This opacity is particularly problematic in a clinical context where understanding the rationale behind a diagnosis is critical (Amann et al., 2020). Moreover, when models provide outcomes that differ from a medical expert’s assessment, the inability to explain the discrepancy undermines trust. Regulatory and ethical considerations further elevate the importance of explainability, with frameworks such as the GDPR emphasizing the need for traceability of decisions (Holzinger et al., 2017). The emergence of Explainable AI techniques—such as SHAP (Lundberg & Lee, 2017), LIME (Ribeiro et al., 2016), and Integrated Gradients (Sundararajan et al., 2017)—offers pathways to demystify model decision-making. These methods can highlight salient features and regions in medical images that most heavily influence predictions. Yet, the effectiveness of these explanations remains insufficiently quantified. Studies have often focused on visually inspecting feature attribution maps rather than rigorously evaluating the quality, faithfulness, robustness, and complexity of the explanations they provide.

This work employs a rigorous pipeline to train and evaluate deep learning models on the HAM10000 dataset. Multiple architectures are explored, including CNNs designed from scratch and well-known networks such as DenseNet, MobileNet, and ResNet. Data augmentation and oversampling address class imbalances and improve model generalization. After achieving strong predictive performance, several xAI techniques are applied to generate post hoc explanations, revealing the image features that guide the model’s predictions. The central contribution is the quantitative evaluation of these explainability techniques. By employing metrics that assess faithfulness, robustness, and complexity, this study establishes measurable standards for comparing and refining xAI methods. Adjusting the parameters and hyperparameters of explanation algorithms and evaluating their performance under these metrics enables data-driven improvements in the quality of explanations.

The methodology involves three stages: (1) model training, (2) explainability generation, and (3) quantitative evaluation. After successfully training the classification models and validating their performance on held-out test sets, xAI techniques are used to generate explanatory maps. A suite of metrics derived from existing xAI literature is then employed to quantitatively assess these explanations. This framework ensures a holistic evaluation of both model accuracy and the explanatory power of the outputs. Results demonstrate that altering hyperparameters of explanation methods can significantly influence their measured quality. Identifying which configurations yield more faithful and robust explanations provides preliminary guidelines for applying xAI in medical diagnostics. While limitations such as data diversity and computational intensity persist, this research offers actionable insights for developing more transparent and clinically acceptable AI tools. Furthermore, the techniques and lessons learned here can be extended to other medical imaging domains.

**Figure 1:**
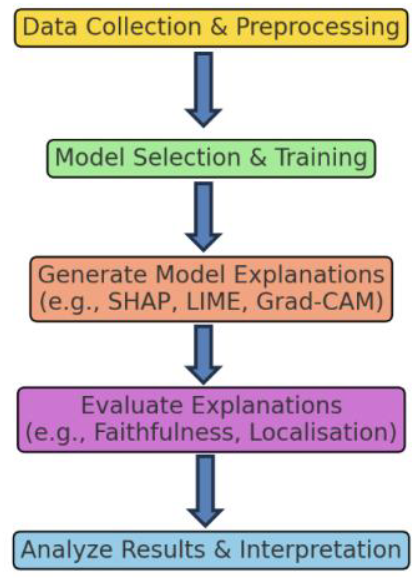
Solution Approach

## 2 Background and Related Work

This section surveys key areas relevant to increasing transparency and reliability: the models and architectures commonly employed for medical image classification, the fundamentals of explainable AI (xAI) methods, and approaches to evaluating explainability—particularly in healthcare contexts.

Deep learning model performance is heavily influenced by hyperparameters such as learning rate, batch size, and number of epochs, as well as by data augmentation techniques and class balancing strategies. The learning rate, for instance, dictates the size of optimization steps toward minimizing loss. Adaptive strategies, such as reducing the rate upon performance plateaus, help fine-tune convergence (Bengio, 2012). Early stopping is often employed to prevent overfitting, a critical concern in medical diagnostics where models must generalize beyond training data. Data augmentation, including rotations, flips, and crops, diversifies the training set and improves generalization (Perez & Wang, 2017). Oversampling techniques—such as random oversampling and SMOTE (Chawla et al., 2002)—address class imbalance, a frequent issue in clinical datasets. Metrics for evaluation, such as precision, recall, F1 score, Matthews Correlation Coefficient (MCC), and AUC-ROC, provide a more nuanced assessment of model performance than accuracy alone, particularly in imbalanced scenarios (Esteva et al., 2017; Chicco & Jurman, 2020; Hicks et al., 2022).

**Figure 2:**
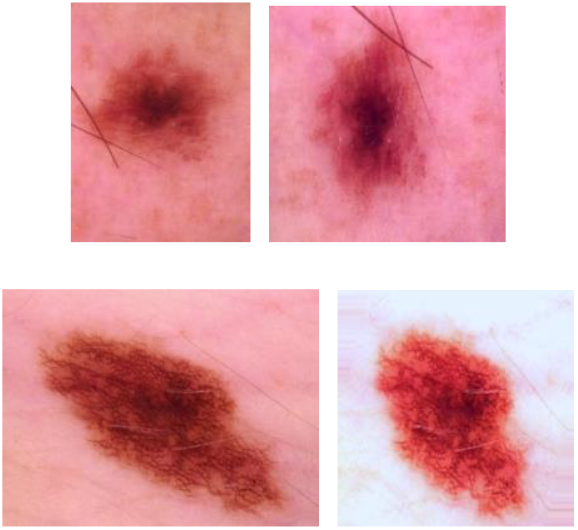
Some Sample Input and Augmented Images

Convolutional Neural Networks (CNNs) efficiently learn hierarchical representations from image data, making them particularly effective for lesion classification tasks. By convolving filters over image pixels, CNNs capture spatial and textural features at multiple scales, enabling reliable feature extraction and classification (Hosny et al., 2018; Alfi et al., 2022; Alkarakatly et al., 2020). Transfer learning leverages pre-trained models—such as DenseNet, ResNet, and MobileNet—trained on large, general-purpose datasets like ImageNet. DenseNet’s dense connectivity mitigates vanishing gradients and promotes feature reuse (Huang et al., 2017). ResNet employs residual blocks to allow training of deeper architectures without performance degradation (He et al., 2016), while MobileNet offers efficiency through depth wise separable convolutions (Howard et al., 2017). Fine-tuning these models to domain-specific tasks, such as skin lesion classification, can significantly reduce training time and computational demands, while improving performance under limited data conditions. Inception architectures (Szegedy et al., 2015) further enhance representational power by integrating multiple convolution filters within a single layer, capturing features at varying spatial scales.

**Figure 3:**
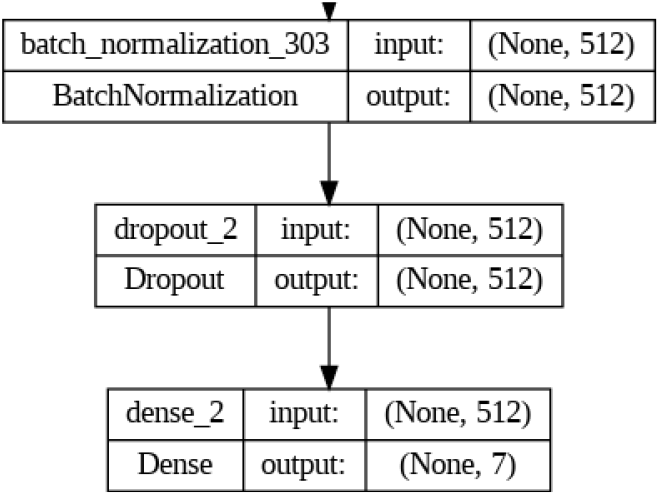
Some of the top layers of a transfer learnt DenseNet121 Model used in this study

Research in healthcare AI has increasingly integrated explainability. Surveys by Wang (2023) underscore the role of uncertainty quantification and xAI in improving model transparency. Other studies have employed LIME, SHAP, and similar methods for early disease detection, enhancing clinical understanding and informing treatment decisions (Magesh et al., 2020; Singh et al., 2020; Shahsavari et al., 2023). Efforts such as those by Pieter Van Molle et al. (2018) and Cristiano Patrício et al. (2023) emphasize explainability in dermatological imaging, facilitating improved clinical decision support. Similar trends appear in cancer classification (Yaqoob et al., 2023), COVID-19 detection (Xu et al., 2021), and lung and breast cancer diagnostics (Sobhan & Mondal, 2021; Binder et al., 2021; Rezazadeh et al., 2022; Ladbury et al., 2022).

**Figure 4:**
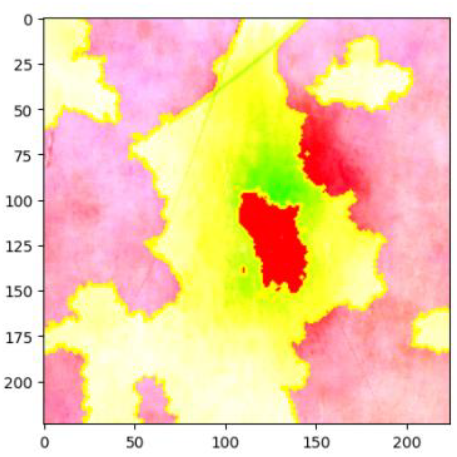
Example of a LIME Explanation. The green area signifies positive relationship with the label and the red signifies a negative on with the particular label under consideration. Axes are pixels, with the image of size

**Figure 5:**
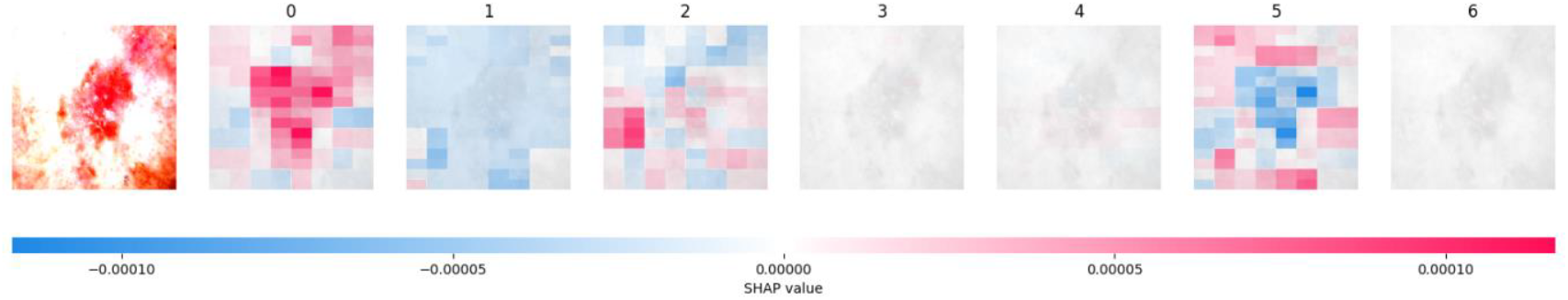
SHAP Example, here red denotes positive attribution towards the label and blue denotes negative. In this correctly classified image (label 0), you can see the features/regions that contributed to that output

Assessing the effectiveness of xAI methods remains a pressing issue. Evaluation strategies range from qualitative approaches—user and expert studies examining clarity, coherence, and narrative quality of explanations—to automated quantitative metrics. Common metrics include:

1. Faithfulness: measures how accurately an explanation reflects the true reasoning process of the AI model. For example, faithfulness correlation evaluates the correlation between importance assigned by the explainer and the actual impact of those features on the model’s predictions (Bhatt et al., 2020).
2. Localization: assesses how well an explanation identifies the relevant parts of the input. Localisation metrics usually rely on comparing segmentation maps with explainer identified image regions for assessing Intersection over Union for instance.
3. Complexity: measures the cognitive load required for a human to understand the explanation. Lower complexity indicates a more interpretable explanation. Rudin (2019) argued for simpler models and explanations, emphasizing that less complex explanations are easier for humans to understand and verify.
4. Plausibility: assesses whether the explanation makes sense to human experts, even if it is not a perfect representation of the model’s internal logic. Kulesza et al. (2015) proposed evaluating explanations based on how plausible they appear to domain experts, regardless of their faithfulness.
5. Robustness: evaluates the stability of explanations under small perturbations of the input. An explanation is robust if small changes in the input do not significantly alter the explanation. Alvarez-Melis and Jaakkola (2018) introduced methods to quantify the robustness of explanations, ensuring they remain consistent under similar conditions.

A key challenge is the lack of standardized benchmarks for explainability, complicating comparisons and reproducibility. While some studies incorporate human-grounded evaluations or employ multiple metrics in tandem (Doshi-Velez & Kim, 2017; Poursabzi-Sangdeh et al., 2018), the field still lacks consensus on the “best” set of metrics.

**Table 1:**
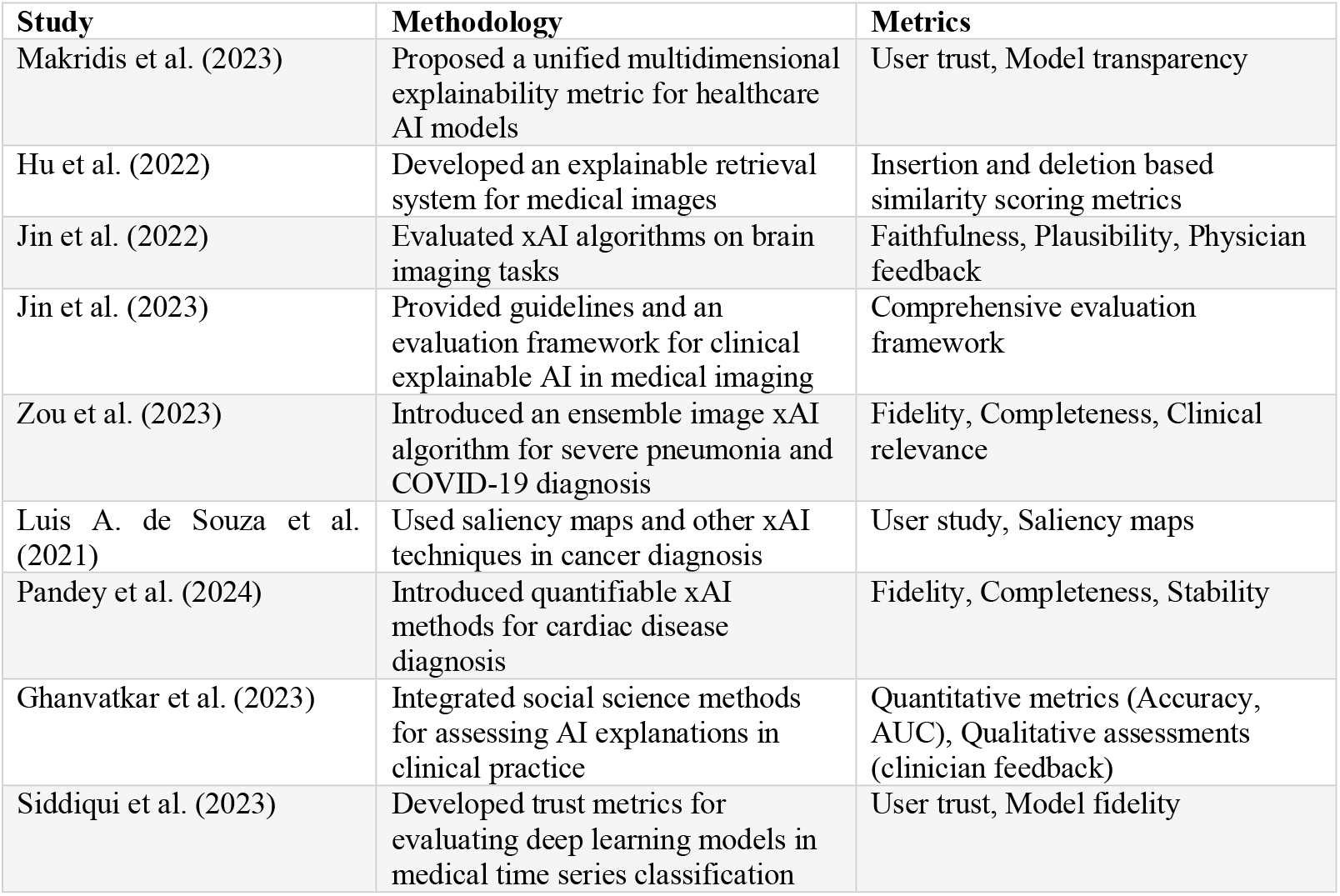
Summary of Key studies in Explainability Evaluation in Healthcare.

| Study | Methodology | Metrics |
| --- | --- | --- |
| Makridis et al. (2023) | Proposed a unified multidimensional explainability metric for healthcare AI models | User trust, Model transparency |
| Hu et al. (2022) | Developed an explainable retrieval system for medical images | Insertion and deletion based similarity scoring metrics |
| Jin et al. (2022) | Evaluated xAI algorithms on brain imaging tasks | Faithfulness, Plausibility, Physician feedback |
| Jin et al. (2023) | Provided guidelines and an evaluation framework for clinical explainable AI in medical imaging | Comprehensive evaluation framework |
| Zou et al. (2023) | Introduced an ensemble image xAI algorithm for severe pneumonia and COVID-19 diagnosis | Fidelity, Completeness, Clinical relevance |
| Luis A. de Souza et al. (2021) | Used saliency maps and other xAI techniques in cancer diagnosis | User study, Saliency maps |
| Pandey et al. (2024) | Introduced quantifiable xAI methods for cardiac disease diagnosis | Fidelity, Completeness, Stability |
| Ghanvatkar et al. (2023) | Integrated social science methods for assessing AI explanations in clinical practice | Quantitative metrics (Accuracy, AUC), Qualitative assessments (clinician feedback) |
| Siddiqui et al. (2023) | Developed trust metrics for evaluating deep learning models in medical time series classification | User trust, Model fidelity |

Recent healthcare-focused studies propose frameworks to evaluate and integrate explainability into clinical workflows. For example, Makridis et al. (2023) introduce a multidimensional evaluation metric, while Hu et al. (2022) apply insertion and deletion-based scoring to gauge explainer quality for medical image retrieval. Jin et al. (2022, 2023) highlight faithfulness, plausibility, and physician feedback as crucial metrics, while Zou et al. (2023) integrate ensemble xAI methods and assess fidelity and clinical relevance. Ensemble approaches, as explored by Pandey et al. (2024) and Ghanvatkar et al. (2023), combine quantitative and qualitative metrics, leveraging social science methodologies and clinical feedback to ensure that explanations are both technically sound and clinically useful. Siddiqui et al. (2023) similarly focus on trust metrics for time-series classification in medical data. From the above, we can note that combining quantitative metrics with qualitative assessments is crucial for a comprehensive evaluation of xAI methods. Equally important is the ensembling of different explanation methods together, an idea employed by at least 2 of the above-mentioned studies. One major issue which is clear from the variety of metrics used in the aforementioned research, is the lack of standardized evaluation frameworks that can be universally applied across different healthcare domains and applications. Ensuring that healthcare professionals trust and understand AI explanations is critical for the successful adoption of xAI methods in practice and so, it is important to integrate end users in the selection of the explanation model.

## 3 Formal Definition and Approach

The central hypothesis is that by systematically comparing the performance of multiple xAI techniques under different hyperparameter settings, it is possible to identify which methods yield the most reliable and interpretable explanations for skin cancer classification. This study aims to address the gap in rigorous, quantitative assessment of xAI methods specifically tailored to medical imaging tasks. Multiple deep learning architectures are employed, including both convolutional neural networks (CNNs) trained from scratch and transfer learning-based models (e.g., DenseNet, ResNet, MobileNet) fine-tuned on the HAM10000 dataset. These models differ in complexity, parameterization, and feature extraction strategies. Each model seeks to minimize classification error and maximize diagnostic accuracy while handling constraints such as limited data and the need for robust generalization. Building upon studies that highlight the importance of explainability in healthcare AI (Wang, 2023; Magesh et al., 2020; Singh et al., 2020; Shahsavari et al., 2023; Pieter Van Molle et al., 2018), the research outlined here seeks to quantitatively evaluate the performance and interpretability of xAI methodologies for skin cancer classification. These efforts align with previous frameworks (e.g., Ladbury et al., 2022) that emphasize model-agnostic and clinically oriented approaches to explainability. The classification task focuses on the multi-class diagnosis of skin lesions using the HAM10000 dataset, which includes seven categories: Actinic keratoses (akiec), Basal cell carcinoma (bcc), Benign keratosis-like lesions (bkl), Dermatofibroma (df), Melanocytic nevi (nv), Melanoma (mel), and Vascular lesions (vasc). Formally, the problem can be expressed as:

- Input: Dermoscopic images

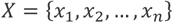

- Output: Corresponding labels

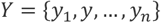

where

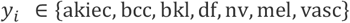

Categorical Cross Entropy Loss:

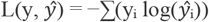

where y is the true label, ŷ is the predicted probability, summed over the number of classes.

Adam combines Adaptive Gradient Descent with Root Mean Squared Propagation. The steps for Adam are as follows:

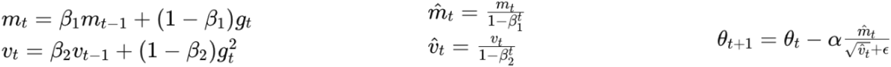

where α is the learning rate, β_1_ and β_2_ are the exponential decays for the first and second moments respectively, m_t_ is the first moment (mean of gradients), t is the time step, ν_t_ is the second moment (variance of gradients), θ_t_ are the model parameters at time t and ε is a small constant to prevent division by zero.

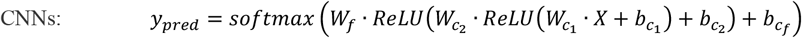

While this is an oversimplification of a more complex CNN, the input image X is passed through two convolutional layers with ReLU activation, followed by a fully connected layer with SoftMax to output class probabilities.

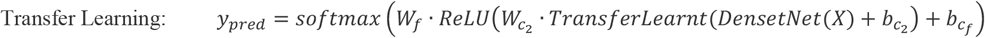

The input image X is processed through a pre-trained transfer learning model, followed by a fully connected layer to output class probabilities. Practically, a lot more top layers would be added onto the model pre-trained on ImageNet weights and then some of the model’s layers would also be unfrozen for further fine tuning.

Post Hoc Explainable AI Methods:

1. <u>SHAP (SHapley Additive exPlanations):</u> Quantifies the contribution of each feature to the final prediction, providing a global view of model behaviour

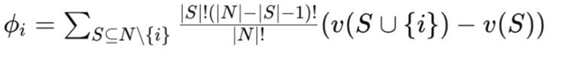

where S is a subset of features, N is the set of all features and v(S) is the prediction when only features in S are present.
2. <u>LIME (Local Interpretable Model-agnostic Explanations):</u> Generates locally faithful explanations by approximating the model locally with an interpretable one

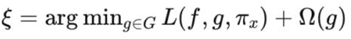

where L is the loss function, f is the black-box model, g is the interpretable model, π is a proximity measure to the instance x, and Ω(g) is a complexity measure.
3. <u>Integrated Gradients:</u> Computes the average gradients of the output with respect to input features, providing insights into feature importance

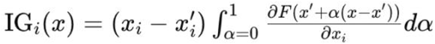

where x is the input, x′ is the baseline, and F is the model. The baseline can be a zero vector, black/white image, a blurred version of the input image or a random/uniform distribution. In this study, a random baseline was used (Fong et al, 2017).

Next, we can look at some of the quantitative metrics commonly used for evaluating the explainability methods:

- <u>Faithfulness</u> is the correlation between importance score & actual impact of features

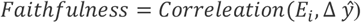

where E_i_ is the score for feature i and Δ ŷ is the change in the prediction when feature i is perturbed or removed.
- <u>Sensitivity</u> evaluates explanation change with input changes, ensuring consistent identification of important features

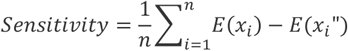

where E(x_i_) is the explanation for input x_i_ and E(x’_i_) is the explanation for a slightly perturbed input x’_i_
- <u>Infidelity</u> is the difference between the explanation and the actual impact of feature perturbations

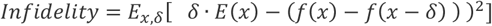

where δ is a perturbation, E(x) is the explanation for input x and f(x) is the model’s prediction for input x.
- <u>Monotonicity</u> assesses if changing the importance score of a feature leads to a consistent change in the prediction

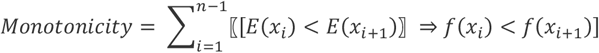

where E(x_i_) is the explanation score, and f(x_i_) is the model prediction for input x_i_
- <u>Sparseness</u> evaluates simplicity by measuring the ratio of features assigned a non-zero importance score.

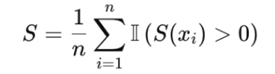

where S(x_i_) is the important score for feature x_i_ and n is the number of features
- <u>Relative Stability</u> measure consistency with respect to input perturbations.

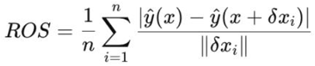

where n is the number of perturbations δx is the magnitude of the perturbation, y is the model output.
- <u>IROF (Input Reduction Output Fidelity)</u> measures how much of the input data can be reduced (removed or masked) while maintaining the model’s original output.

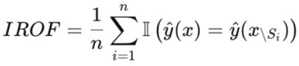

where n is the number of samples, x is input with features S removed and y is the output of the model.

## 4 Methodology

This section provides a comprehensive overview of the methodology used to address the problem of developing and evaluating explainable models for the diagnosis of skin cancer using the HAM10000 dataset. The methodology involves data preprocessing, classification model development, training, evaluation, and the application of explainable AI (xAI) methods to quantify and compare model interpretability.

The HAM10000 dataset comprises 10,015 dermoscopic images, categorized into seven classes: actinic keratoses, basal cell carcinoma, benign keratosis-like lesions, dermatofibroma, melanocytic nevi, melanoma, and vascular lesions (Tschandl et al., 2018). This dataset was chosen due to its class diversity, clinical relevance and the amount of previous classification research available on it for reference and comparison. The dataset was divided into training and validation sets using an 75-25 split, with an additional 1500 test images used to finally test the generalizability of the classifiers. Stratified sampling was employed to maintain class distribution across all sets. To enhance model robustness, image augmentation techniques such as channel and brightness shifts, shears, random rotations, flips and zooms were applied to the training set (Perez & Wang, 2017). Oversampling was used to reduce class imbalance and rates were based on previous research suggesting a max 300% increase in the smaller classes. Image pixel values were normalized to improve model convergence rates.

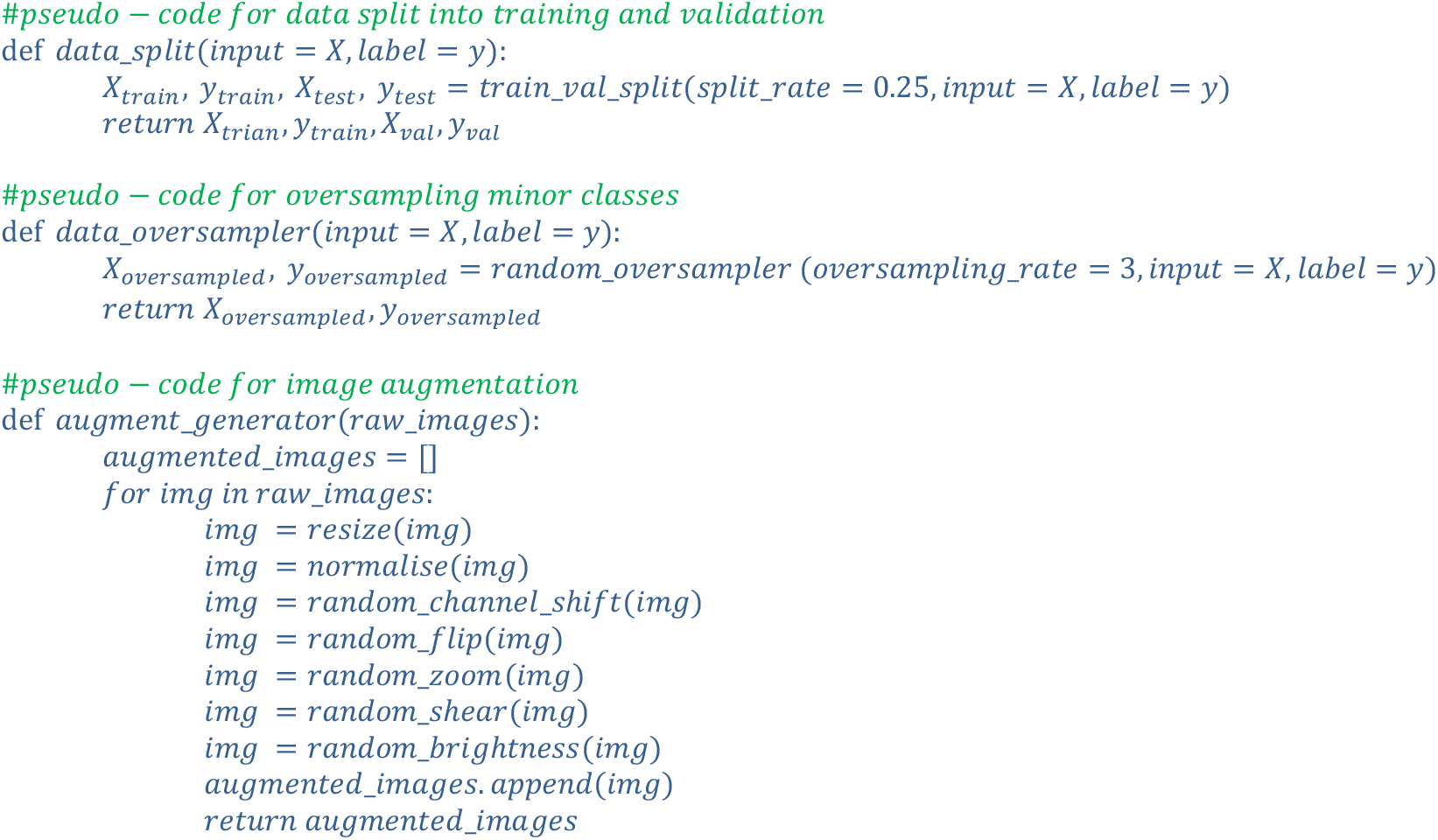

CNNs were built from scratch with varying architectures, including different numbers of convolutional layers, kernel sizes, and activation functions. The architectures were optimized through hyperparameter tuning. Pre-trained models such as DenseNet, ResNet, and MobileNet were fine-tuned on the HAM10000 dataset. These models were chosen for their proven efficacy in image classification tasks. Loss Function and Optimization: Categorical cross-entropy was chosen due to its suitability for multi-class classification problems. The Adam optimizer was used over Stochastic Gradient Descent (SGD) because of its adaptive learning rate capabilities, which typically result in faster convergence (Kingma & Ba, 2014). While SGD can perform better, it can be a lot slower to converge. Reduce on plateau was used to reduce the learning rate when the validation loss plateaued, allowing the models to converge more smoothly. Experiments were conducted with batch sizes of 8, 32, 64, and 128. The number of epochs varied between 50 and 200, depending on the convergence behaviour observed during training.

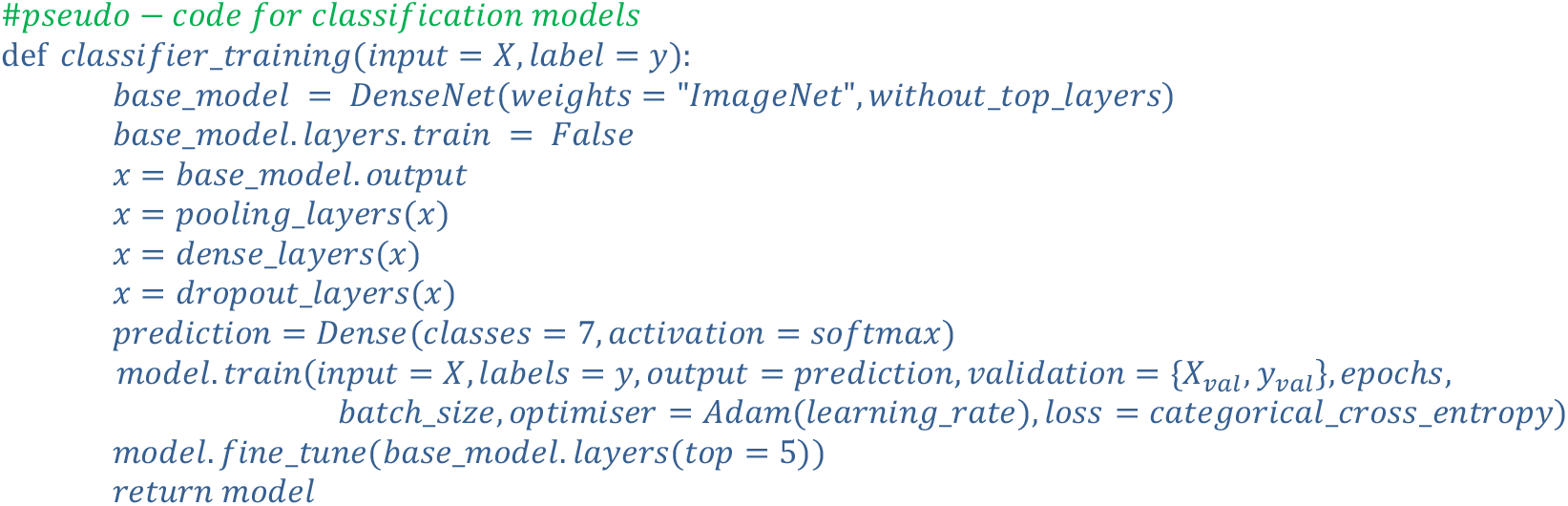

Post hoc explainability methods such as Integrated Gradients, LIME and SHAP were applied to selected models to generate explanations for their predictions. These perturbations and saliency-based methods help in understanding the contribution of each pixel and feature to the decision of the model.

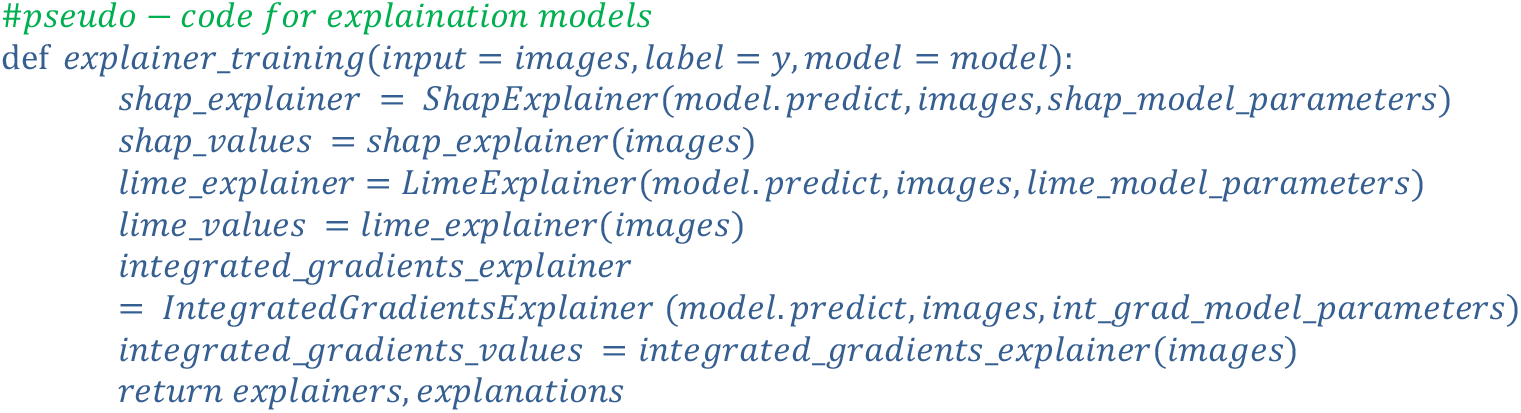

Text based explanations and their accompanying evaluation using methods such as METEOR (Metric for Evaluation of Translation with Explicit Ordering), ROUGE (Recall-Oriented Understudy for Gisting Evaluation) and BLEU (Bilingual Understudy Evaluation) has been seen several times before (Patricio et al., 2023). This paper focuses instead on image-based explanation methods such as Integrated Gradients, LIME and SHAP and evaluates their performance quantitatively. Classification Models were evaluated using standard metrics such as accuracy, MCC, recall, precision, AUC and F1 score. These metrics comprehensively assess the performance of the model, particularly in the context of imbalanced datasets. The effectiveness of xAI methods was quantified using metrics such as faithfulness correlation, sensitivity, stability, etc on different variations of the explainers for comparisons. These metrics help in assessing how well the explanations align with the classification model’s actual resolution process.

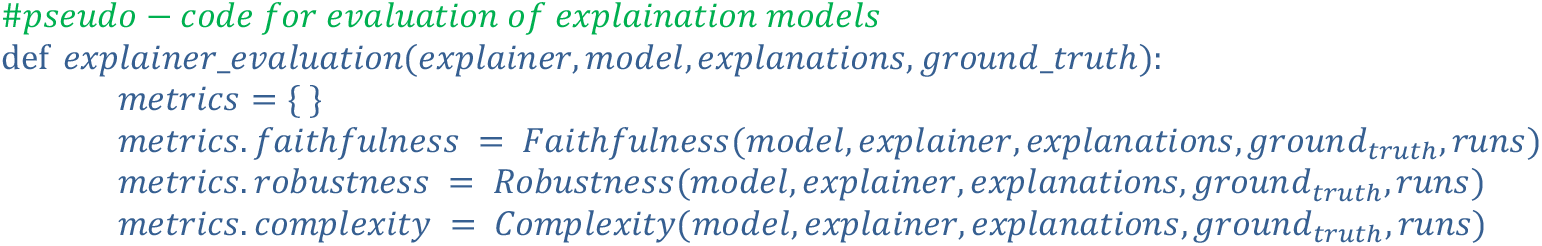
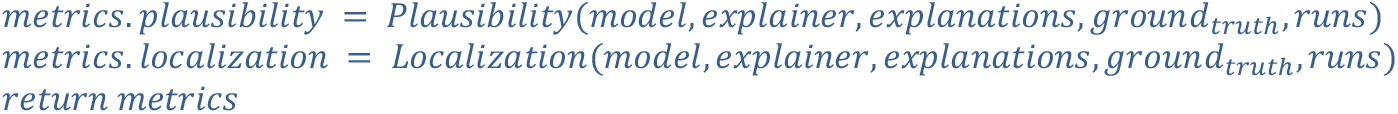

Experiments were conducted on a Google Cloud Workbench with 1 NVIDIA L4 GPU along with 4 2-core vCPUs with 16 GB memory to accelerate the training and evaluation processes. The models were implemented using TensorFlow and Keras frameworks. Code was written from scratch as well as adapted from publicly available repositories as needed, with modifications made to suit the specific requirements of this project. Appropriate references to original authors were provided where necessary in the codebase. The code and trained models, along with documentation, were made available on GitHub (Sangwan 2024).

## 5 Results

The key findings are organized into sections focusing on data analysis, impact of oversampling and augmentation techniques, performance of the trained classification models, and evaluation of explanation methods. This analysis allows for a thorough understanding of the success and limitations of models and methods applied in this study.

### 5.1 Data Preprocessing

The dataset, HAM10000, initially exhibited significant class imbalance, with some classes being underrepresented. To mitigate this, oversampling techniques were employed, particularly focusing on randomly replicating instances from the minority classes. The original dataset distribution revealed a significant skew towards certain classes, potentially leading to biased model training. This imbalance posed a challenge in accurately predicting the underrepresented classes. After applying random oversampling, the dataset was more balanced, ensuring that minority classes were adequately represented during the training process.

**Figure 6:**
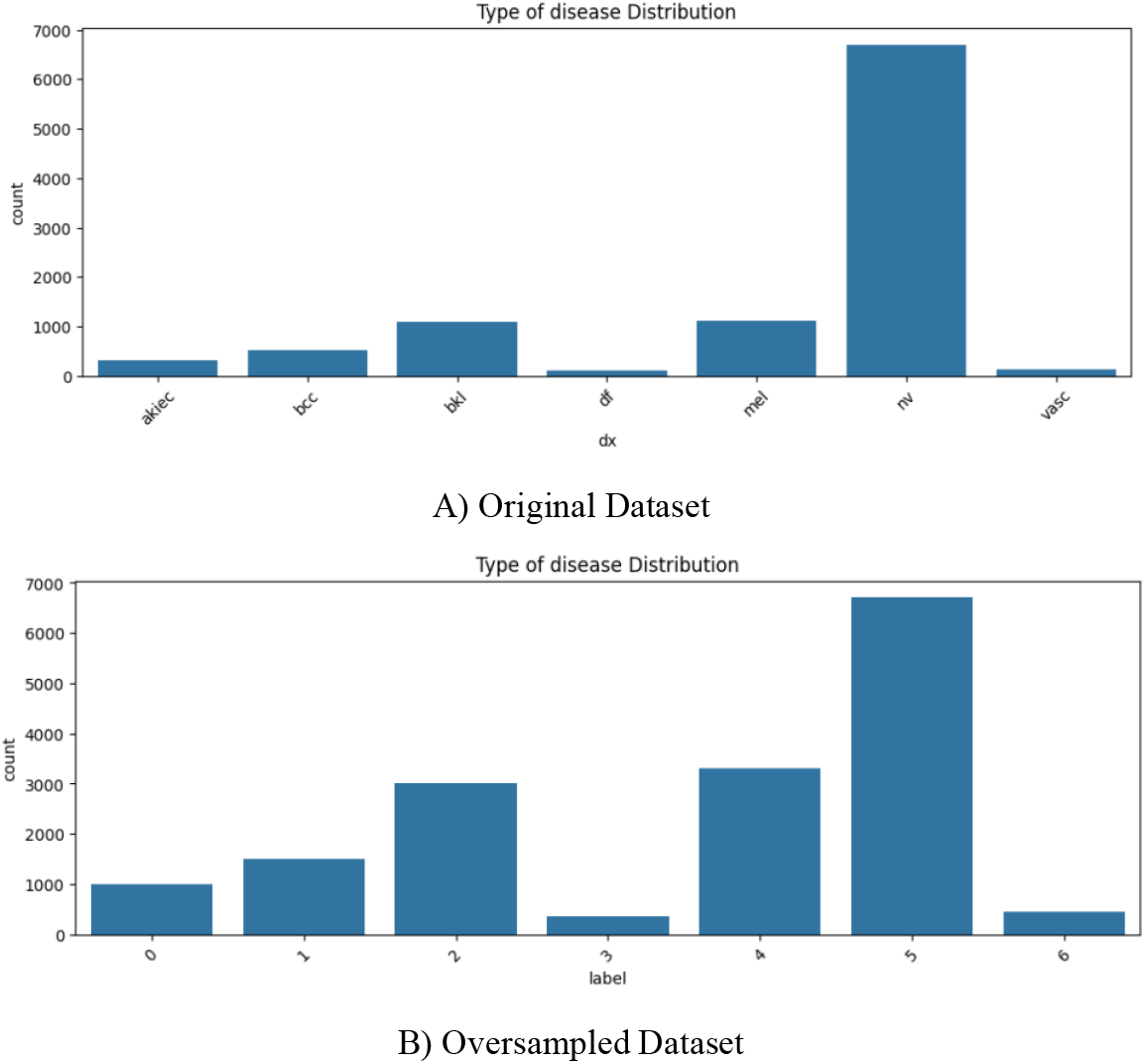
Comparing the distribution of the original vs the oversampled dataset

Augmentation involved generating multiple variations of input images through random transformations, such as rotations, translations, flips, and zooms. This approach increased the effective size of the training dataset while exposing the model to a wider variety of scenarios. A new random augmentation is applied to every image in each epoch. The two figures below showcase the augmentations. The first figure shows how the same image is augmented slightly differently in every epoch, thereby giving the model a bit more information. The second figure showcases how the augmentation is applied to a batch of images (batch size of 32 shown in the example).

**Figure 7:**
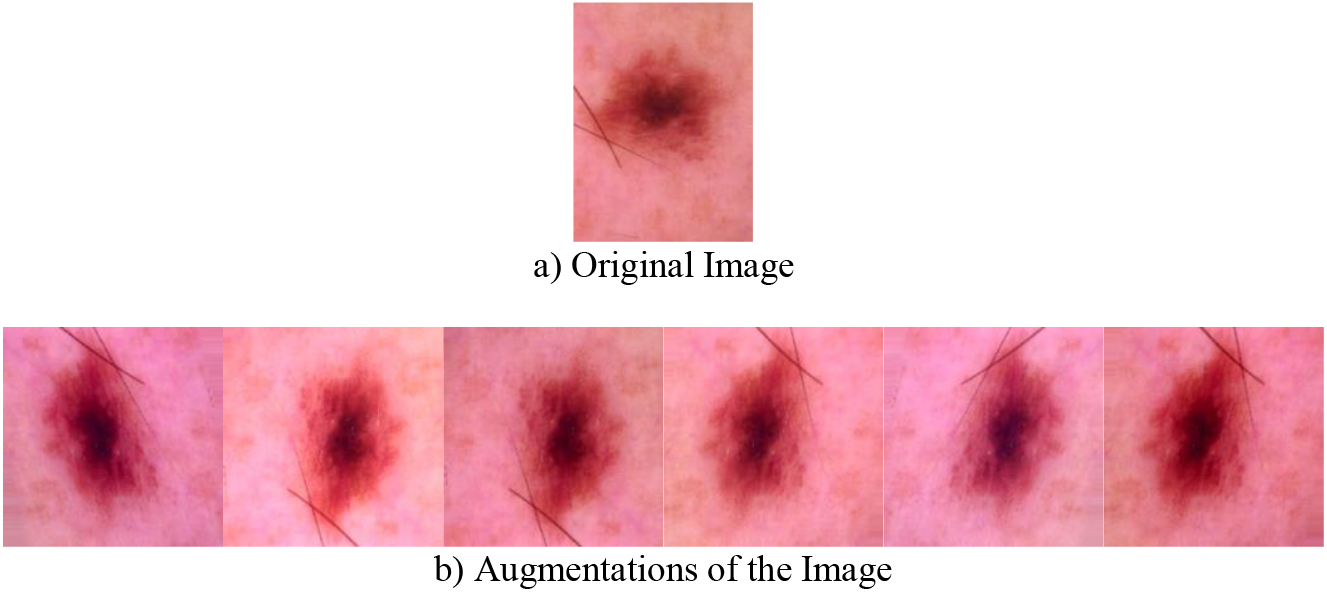
Example of different augmentations of the same image over different epochs.

**Figure 8:**
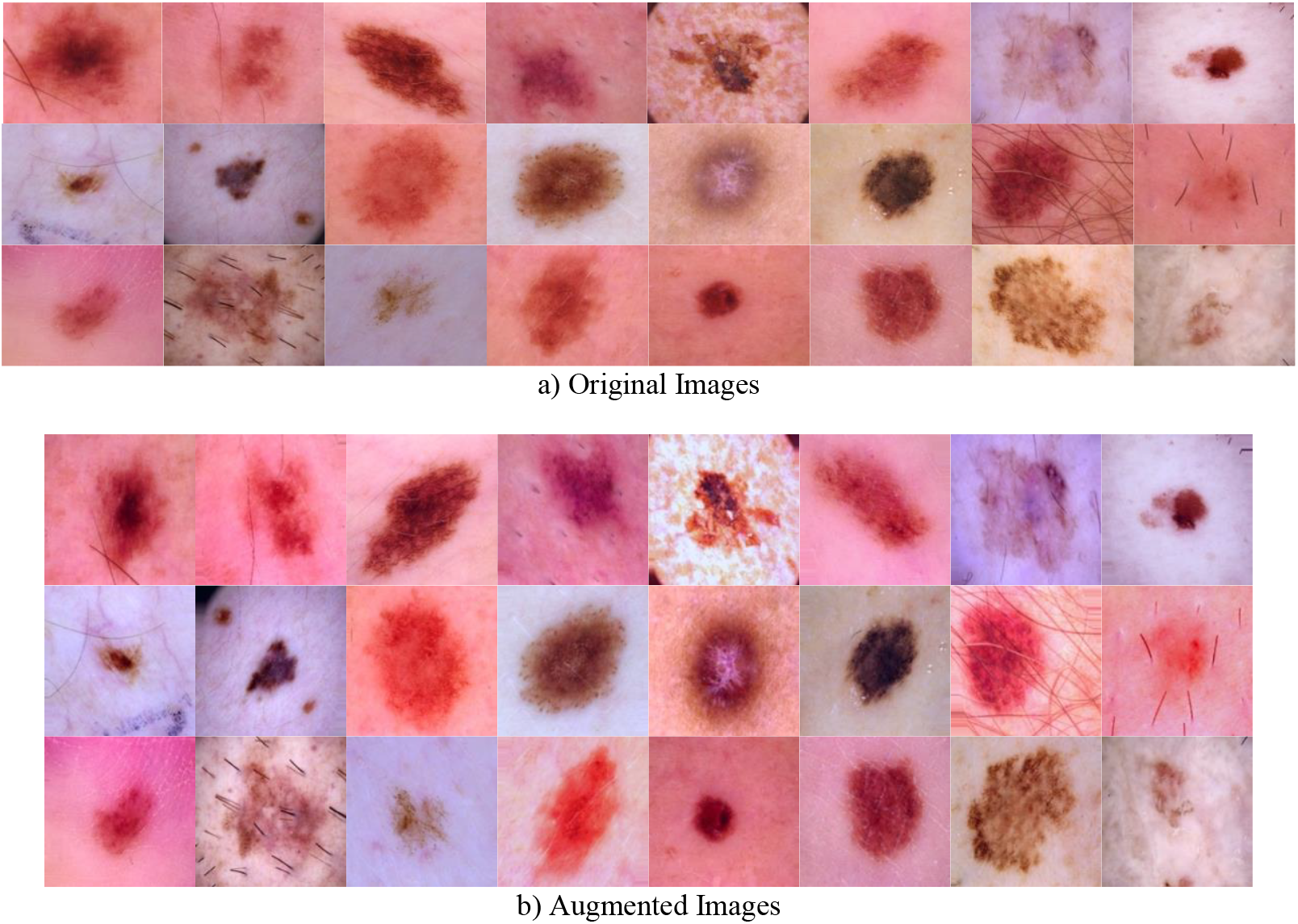
Example of a batch of images and their augmentations for a single epoch. New augmentations each epoch

### 5.2 Classification models

The study developed and tested several classification models, each trained and validated using the augmented and oversampled dataset. The models included a custom-built CNN as well as pre-trained models such as DenseNet and MobileNet, fine-tuned through transfer learning. The performance of these models was evaluated using a set of metrics, with the results shown below in comparison with other state-of-the-art research in this field. Please note that the results are from the validation set. Tested on a completely unseen set from a different source of images, the model achieved accuracy, precision and recall values of around 80%.

**Table 2:**
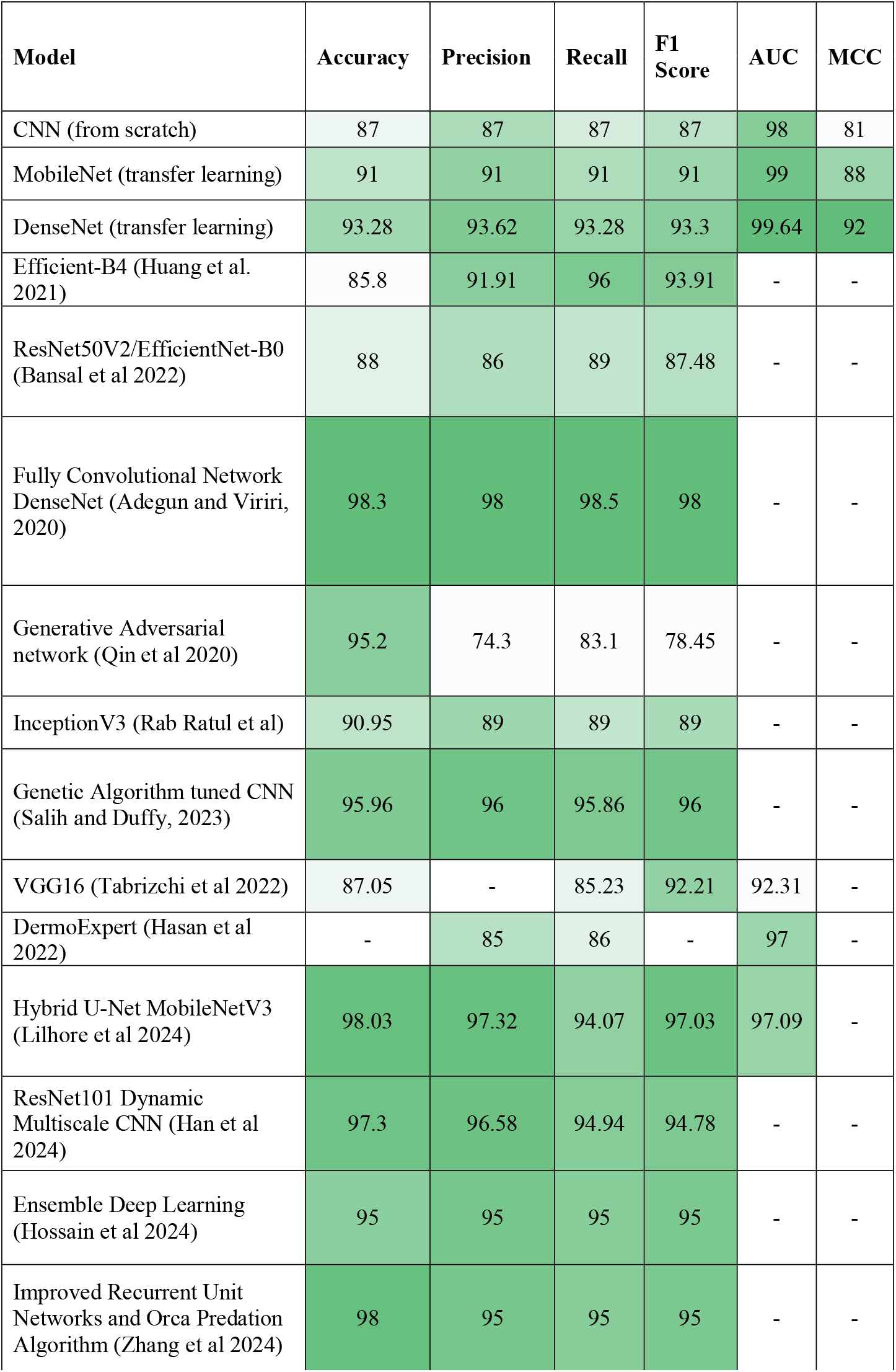
Comparing the trained classification models’ validation metrics with state-of-the-art methods.

| **Model** | **Accuracy** | **Precision** | **Recall** | **F1 Score** | **AUC** | **MCC** |
| --- | --- | --- | --- | --- | --- | --- |
| CNN (from scratch) | 87 | 87 | 87 | 87 | 98 | 81 |
| MobileNet (transfer learning) | 91 | 91 | 91 | 91 | 99 | 88 |
| DenseNet (transfer learning) | 93.28 | 93.62 | 93.28 | 93.3 | 99.64 | 92 |
| Efficient-B4 (Huang et al. 2021) | 85.8 | 91.91 | 96 | 93.91 | - | - |
| ResNet50V2/EfficientNet-B0 (Bansal et al 2022) | 88 | 86 | 89 | 87.48 | - | - |
| Fully Convolutional Network DenseNet (Adegun and Viriri, 2020) | 98.3 | 98 | 98.5 | 98 | - | - |
| Generative Adversarial network (Qin et al 2020) | 95.2 | 74.3 | 83.1 | 78.45 | - | - |
| InceptionV3 (Rab Ratul et al) | 90.95 | 89 | 89 | 89 | - | - |
| Genetic Algorithm tuned CNN (Salih and Duffy, 2023) | 95.96 | 96 | 95.86 | 96 | - | - |
| VGG16 (Tabrizchi et al 2022) | 87.05 | - | 85.23 | 92.21 | 92.31 | - |
| DermoExpert (Hasan et al 2022) | - | 85 | 86 | - | 97 | - |
| Hybrid U-Net MobileNetV3 (Lilhore et al 2024) | 98.03 | 97.32 | 94.07 | 97.03 | 97.09 | - |
| ResNet101 Dynamic Multiscale CNN (Han et al 2024) | 97.3 | 96.58 | 94.94 | 94.78 | - | - |
| Ensemble Deep Learning (Hossain et al 2024) | 95 | 95 | 95 | 95 | - | - |
| Improved Recurrent Unit Networks and Orca Predation Algorithm (Zhang et al 2024) | 98 | 95 | 95 | 95 | - | - |

The figures below show a more detailed view of the models’ performance, showing some examples of the class wise metrics, confusion matrices and training history.

**Figure 9:**
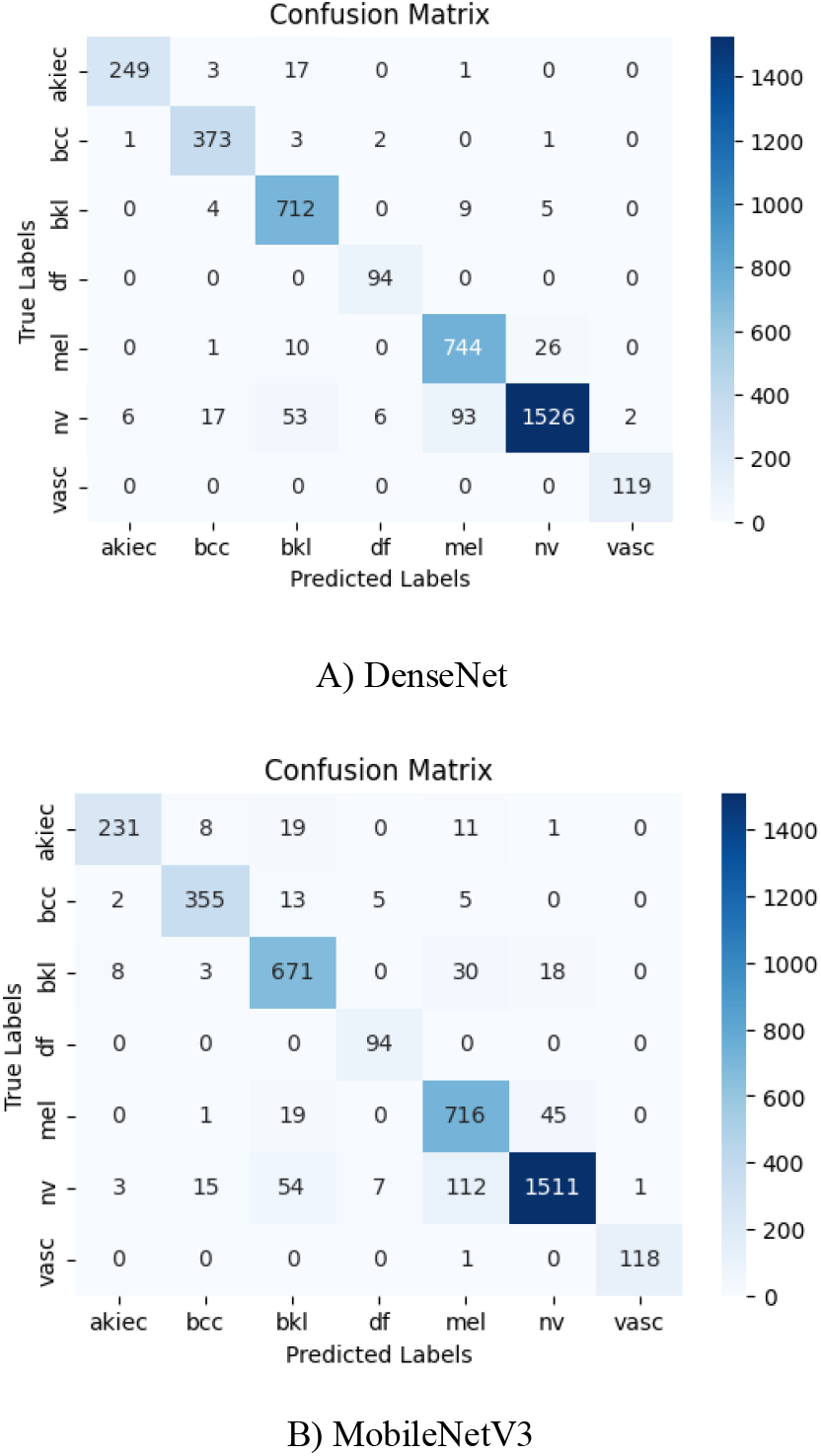
Confusion Matrices on Validation Data

**Table 3:**
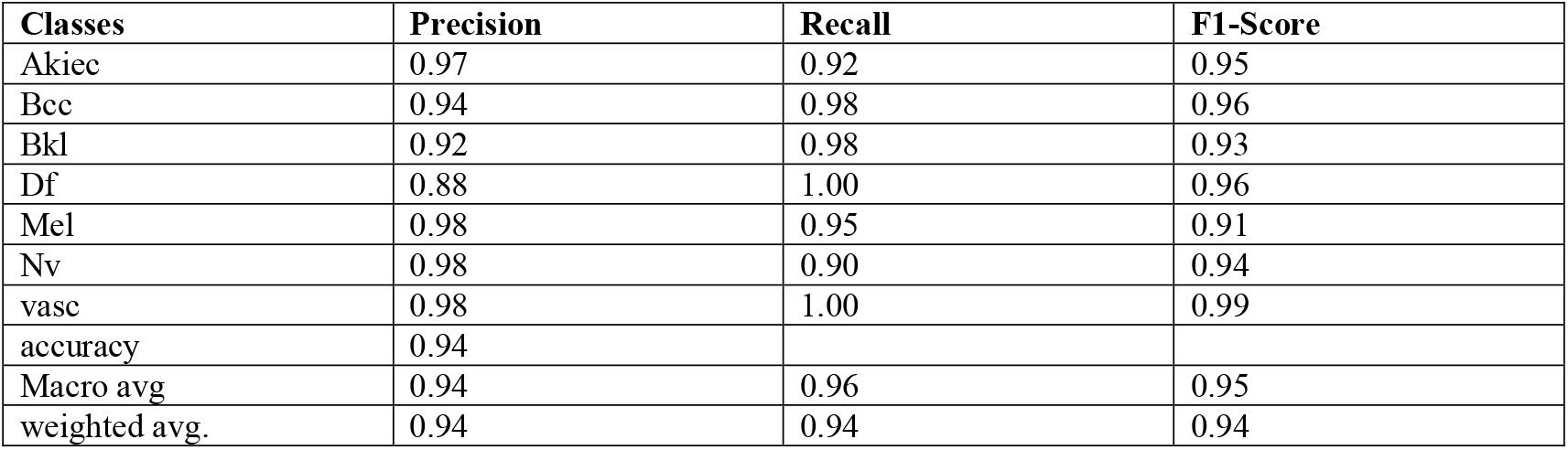
Class-wise metrics example (DenseNet121)

| Classes | Precision | Recall | F1-Score |
| --- | --- | --- | --- |
| Akiec | 0.97 | 0.92 | 0.95 |
| Bcc | 0.94 | 0.98 | 0.96 |
| Bkl | 0.92 | 0.98 | 0.93 |
| Df | 0.88 | 1.00 | 0.96 |
| Mel | 0.98 | 0.95 | 0.91 |
| Nv | 0.98 | 0.90 | 0.94 |
| vasc | 0.98 | 1.00 | 0.99 |
| accuracy | 0.94 |  |  |
| Macro avg | 0.94 | 0.96 | 0.95 |
| weighted avg. | 0.94 | 0.94 | 0.94 |

**Figure 10:**
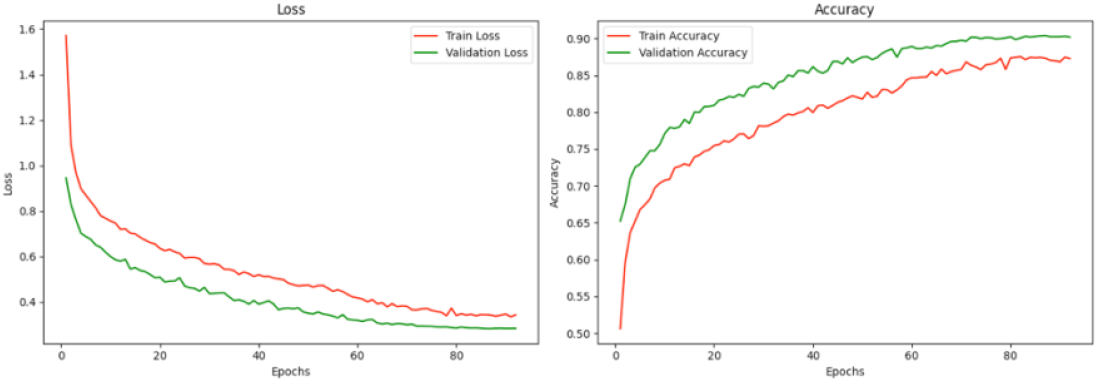
Training History Example (MobileNetV3)

### 5.3 Explanations

Post hoc explanation methods, including Integrated Gradients, LIME and SHAP were applied to the predictions made by the classification models. These methods provided insights into which features contributed to the models’ decisions. Some examples of this implementation are shown below.

**Figure 11:**
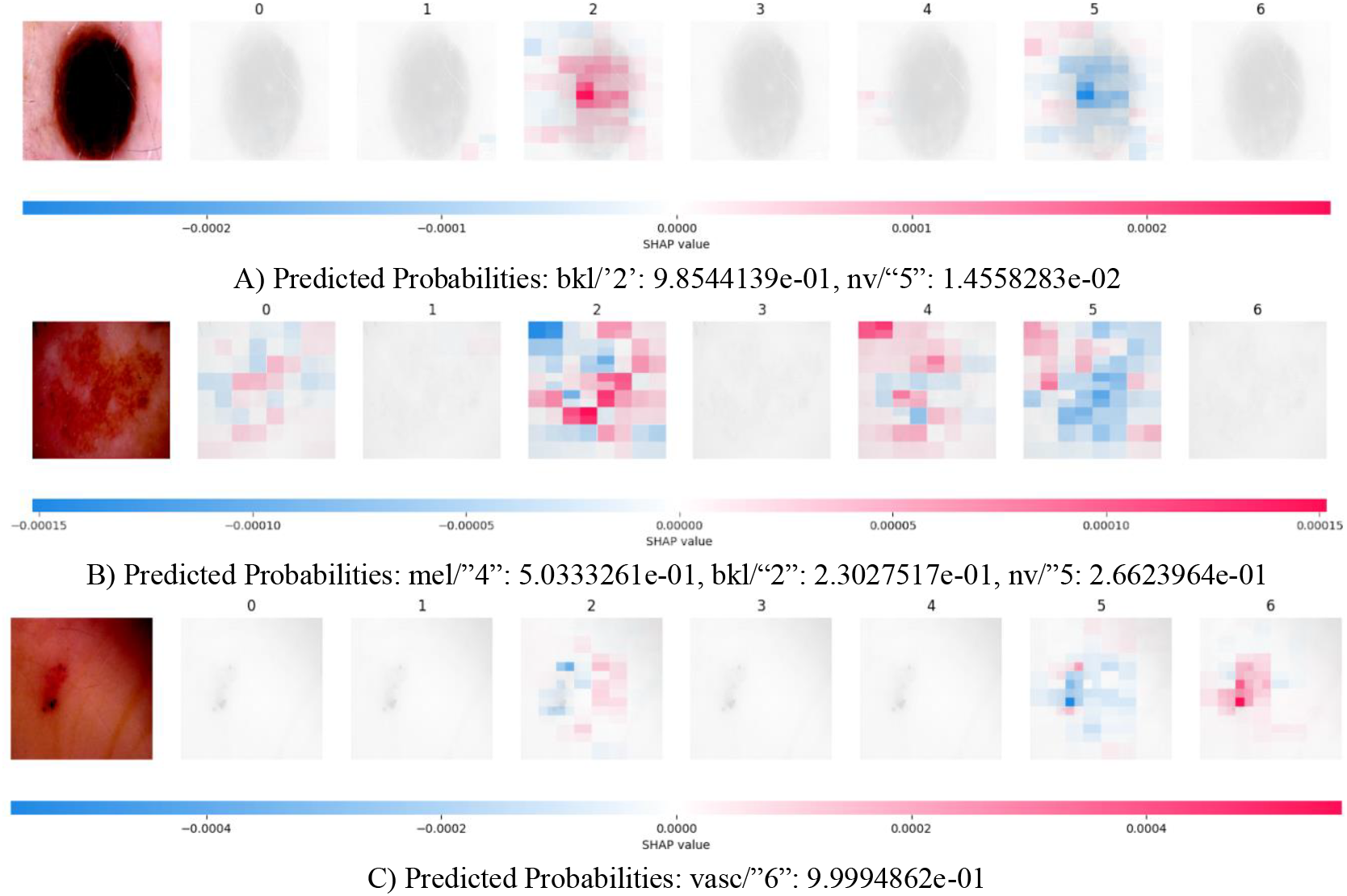
Image on the left and the accompanying map of SHAP values for each label sequentially following it.

**Figure 12:**
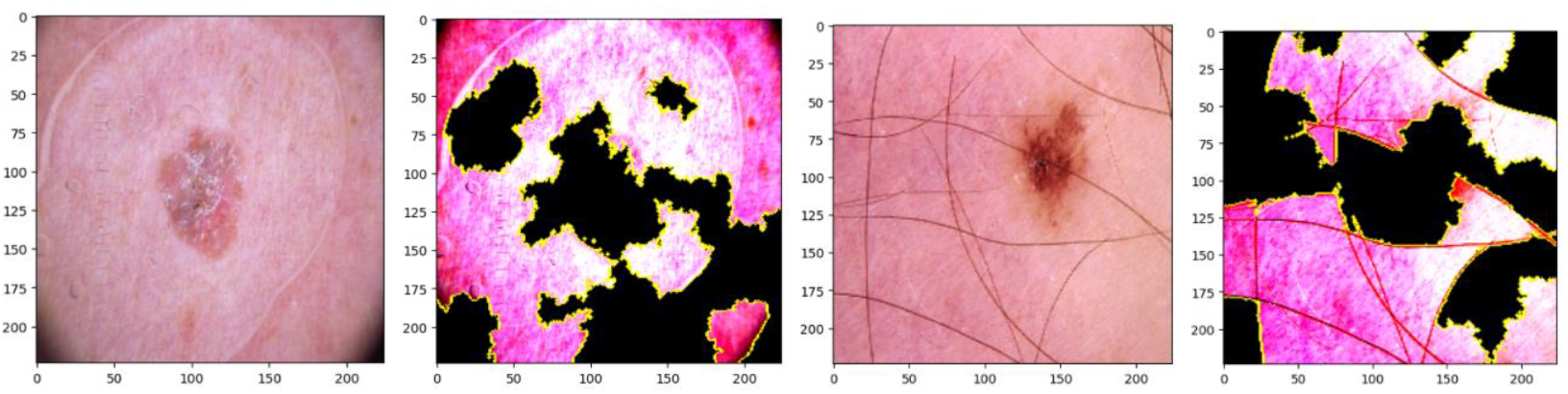
Some examples of lesions and associated masks from LIME, showing the important features. All images are of the top label output of the model.\

**Figure 13:**
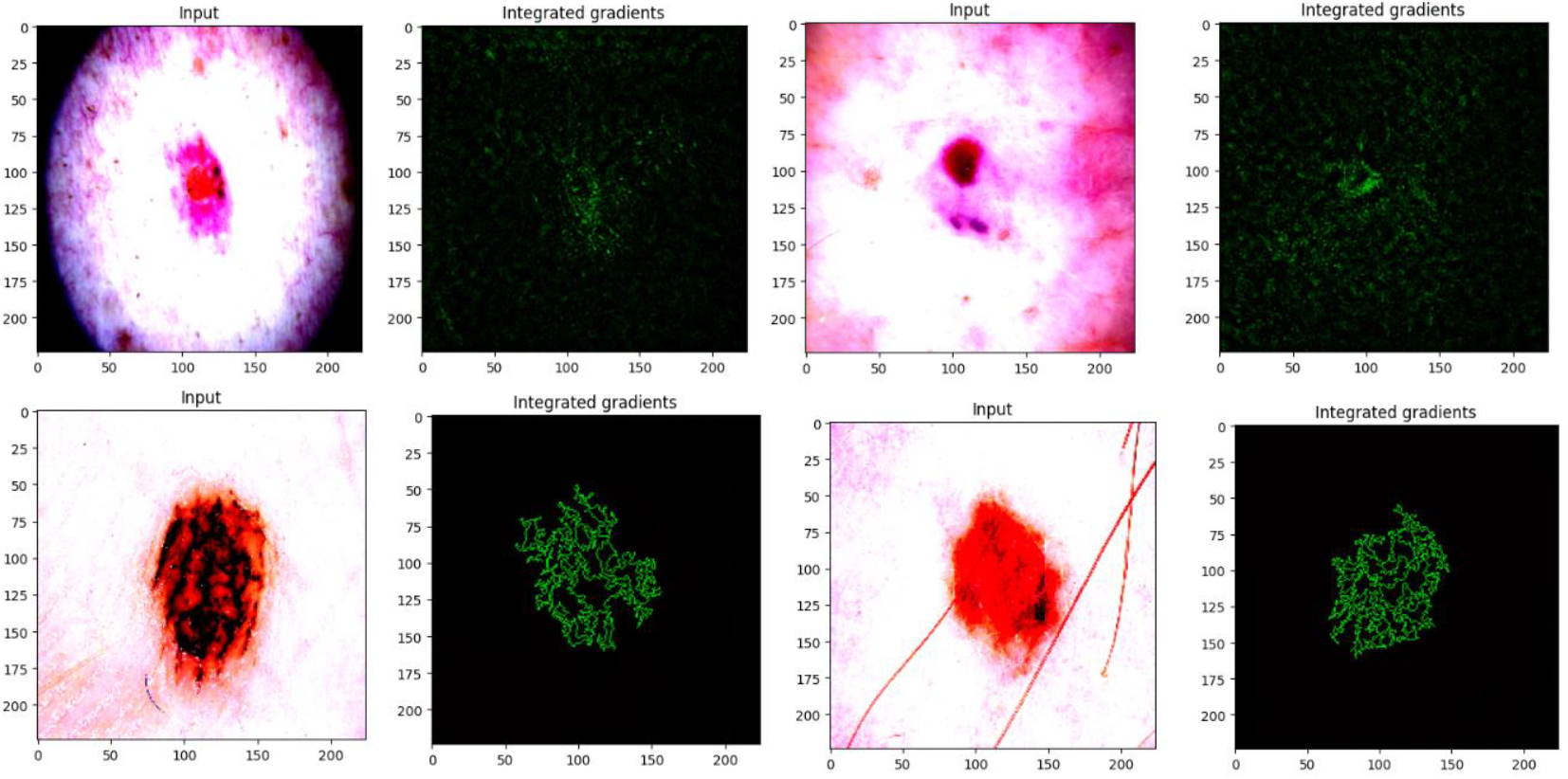
Some examples of Integrated Gradients showing relevant important features for the prediction.

### 5.4 Explanation Evaluations

**Table 4:**
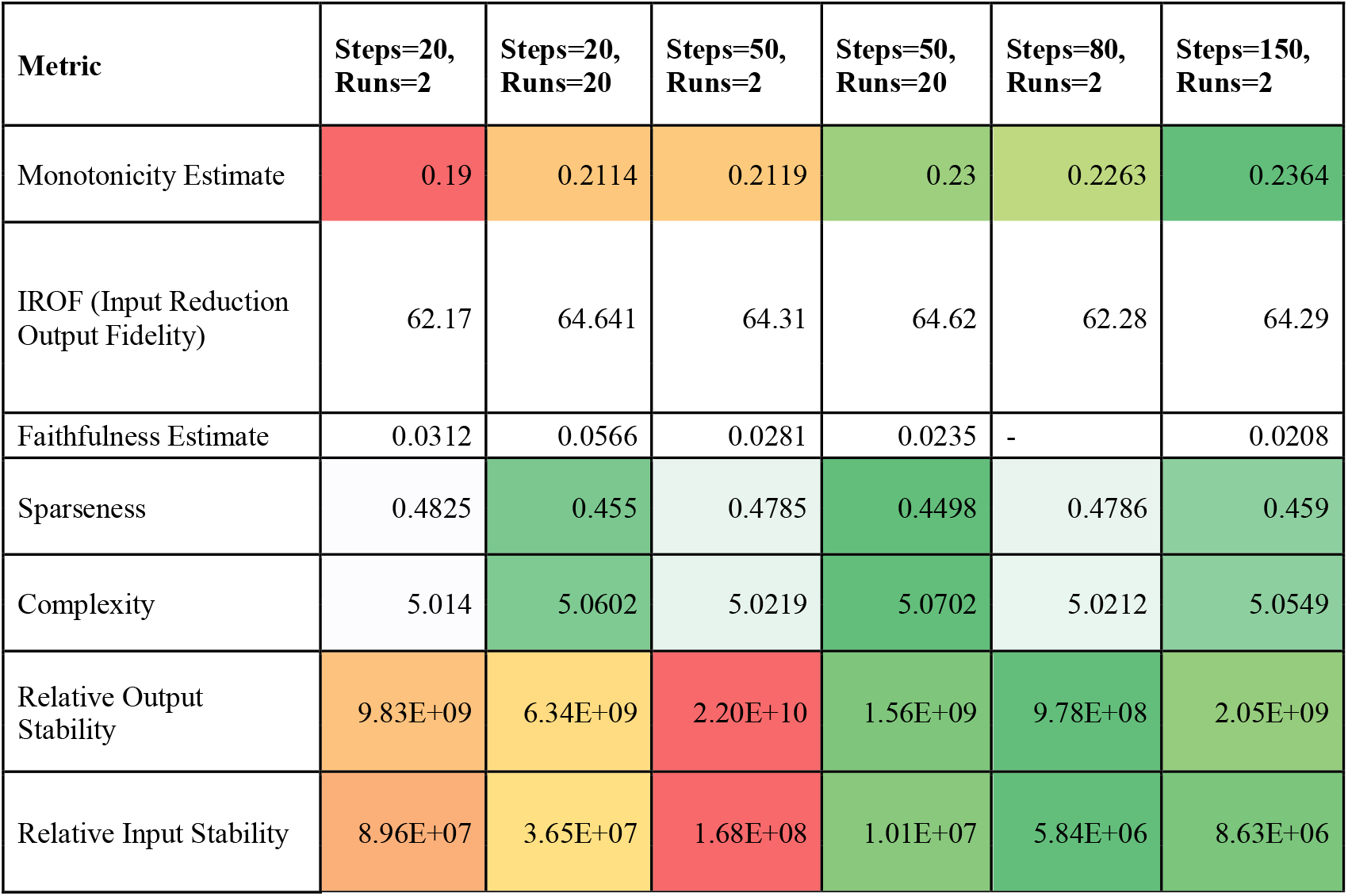
Evaluation of integrated gradients under different hyperparameters.

| Metric | Steps=20,<br>Runs=2 | Steps=20,<br>Runs=20 | Steps=50,<br>Runs=2 | Steps=50,<br>Runs=20 | Steps=80,<br>Runs=2 | Steps=150,<br>Runs=2 |
| --- | --- | --- | --- | --- | --- | --- |
| Monotonicity Estimate | 0.19 | 0.2114 | 0.2119 | 0.23 | 0.2263 | 0.2364 |
| IROF (Input Reduction<br>Output Fidelity) | 62.17 | 64.641 | 64.31 | 64.62 | 62.28 | 64.29 |
| Faithfulness Estimate | 0.0312 | 0.0566 | 0.0281 | 0.0235 | - | 0.0208 |
| Sparseness | 0.4825 | 0.455 | 0.4785 | 0.4498 | 0.4786 | 0.459 |
| Complexity | 5.014 | 5.0602 | 5.0219 | 5.0702 | 5.0212 | 5.0549 |
| Relative Output<br>Stability | 9.83E+09 | 6.34E+09 | 2.20E+10 | 1.56E+09 | 9.78E+08 | 2.05E+09 |
| Relative Input Stability | 8.96E+07 | 3.65E+07 | 1.68E+08 | 1.01E+07 | 5.84E+06 | 8.63E+06 |

All metrics were calculated and averaged over the same batch of 32 lesion images. Due to the computing/time constraints involved, a more involved analysis with further variation in the hyperparameters and additional explainers could not be conducted. But there are still some preliminary insights to be gained from the data above. For example, both relative input and output stability go down as the number of runs are increased. This makes sense since the explainers can average out unimportant noisy features over multiple runs, leading to a better/lower stability score. On the other hand, a clear relationship between number of steps in integrated gradients and stability is not as apparent, although there is an improvement in stability as steps increase. This could possibly be due to the local ‘stability’ minima lying between some values. Of course, the lack of enough data is another possibility for the lack of an apparent relationship over this variable. Sparseness (lower is better) corroborates the insights from stability, suggesting that as more runs are allowed and more parameters made available to increase explainer complexity, it homes in on the most important features.

Complexity seems to increase as the number of steps, or the number of runs is increased. This could suggest the use of this metric to avoid overfitting due to larger hyperparameters that increase model complexity, but only in tandem with another metric that makes sure the explainer isn’t underfit to the model (an explainer that doesn’t do anything would achieve a complexity of zero for example). Monotonicity (higher is better) is another metric that improves as steps or runs are increased. Faithfulness Estimate and IROF (Input Reduction Output Fidelity) are both metrics that fluctuate over the tested space of hyperparameters, making it hard to gain insights from them. It is not clear whether this is an issue with sample space under which they were tested, an issue with the implementation of the calculation itself or whether these metrics might not be suitable for this specific task. It is worth noting that there were also other tested metrics, such as monotonicity, sufficiency and completeness that gave the same results for all combinations (All True/All False for instance) and were therefore not included in the table for differentiation purposes. From this analysis, two things become clear: there are clearly insights to be drawn as to which explainer can perform best by analysing them under different metrics but work still needs to be done to figure out which metrics to use and how to use them in tandem.

## 7 Conclusion

This study addressed the critical challenge of evaluating explainability in AI-driven medical diagnostics, specifically within the context of class-imbalanced skin cancer detection. By employing state-of-the-art convolutional neural networks combined with transfer learning (e.g., DenseNet, MobileNet) and leveraging advanced post hoc explainability methods (e.g., Integrated Gradients, Grad-CAM), the work sought to enhance the interpretability and trustworthiness of automated lesion classification models. The findings demonstrated both the potential and complexity of current xAI techniques in medical applications. While certain evaluation metrics indicated correlations with hyperparameter adjustments, these relationships were not universally consistent, complicating the development of a standardized framework for explainability assessment. The results suggest that one-size-fits-all metrics may not suffice; instead, problem-specific frameworks or the integration of explainability-driven metrics directly into the training process may yield more interpretable and reliable models. For example, incorporating faithfulness or robustness metrics during model optimization could steer learning toward more clinically meaningful explanations rather than relying solely on post hoc evaluation.

Despite these promising insights, several limitations must be noted. First, the reliance on pixel-based explanations may not resonate with clinical reasoning, as they do not directly translate to the semantic features a dermatologist or radiologist would find meaningful. Future research could employ segmentation-based approaches or concept-driven explanations that align more closely with the diagnostic criteria used in practice. Additionally, user studies involving healthcare professionals and the integration of domain-specific knowledge—such as the ABCDE criteria for melanoma—could further ensure that the explanations address genuine clinical needs. Pre-processing measures, like removing image artifacts (e.g., hair), might also refine the clarity of the explanations. Finally, generalizability remains a key area for future work. Testing on diverse datasets and employing real-time, continuous learning models can validate robustness and adaptability. Combining both qualitative expert feedback and quantitative performance metrics can drive a richer understanding of how explainability supports clinical decision-making. In this way, future research can advance beyond pixel-level interpretations toward clinically actionable, concept-based frameworks that strengthen trust, improve patient outcomes, and facilitate the integration of AI into standard medical practice.

## Data Availability

All data produced in the present study are available upon reasonable request to the authors. Some of the code and data is also available at https://github.com/hardikSangwan/thesis_diagnostics_skin

## References

A. A. Adegun and S. Viriri, “FCN-Based DenseNet Framework for Automated Detection and Classification of Skin Lesions in Dermoscopy Images,” in IEEE Access, vol. 8, pp. 150377–150396, 2020, doi: 10.1109/ACCESS.2020.3016651.

Alfi, I.A.; Rahman, M.M.; Shorfuzzaman, M.; Nazir, A. A Non-Invasive Interpretable Diagnosis of Melanoma Skin Cancer Using Deep Learning and Ensemble Stacking of Machine Learning Models. Diagnostics 2022, 12, 726. 10.3390/diagnostics12030726

Ali, A.-R., Li, J. and O’Shea, S.J. (2020). Towards the automatic detection of skin lesion shape asymmetry, color variegation and diameter in dermoscopic images. PLOS ONE, 15(6), p.e0234352. doi:10.1371/journal.pone.0234352.

Alvarez-Melis, David and Jaakkola Tommi S.. “On the Robustness of Interpretability Methods.” (2018)

Amann, J., Blasimme, A., Vayena, E. et al. Explainability for artificial intelligence in healthcare: a multidisciplinary perspective. BMC Med Inform Decis Mak 20, 310 (2020). 10.1186/s12911-020-01332-6

Australian Institute of Health and Welfare (2023). Cancer Data in Australia, Overview of Cancer in Australia, 2023. [online]. Available at: https://www.aihw.gov.au/reports/cancer/cancer-data-in-australia/contents/overview-of-cancer-in-australia-2023

Binder, A., Bockmayr, M., Hägele, M. et al. Morphological and molecular breast cancer profiling through explainable machine learning. Nat Mach Intell 3, 355–366 (2021). 10.1038/s42256-021-00303-4

Bhatt, U., Adrian Weller, and José M. F. Moura. 2021. Evaluating and aggregating feature-based model explanations. In Proceedings of the Twenty-Ninth International Joint Conference on Artificial Intelligence (IJCAI’20). Article 417, 3016–3022.

Brinker TJ, Hekler A, Enk AH, Klode J, Hauschild A, Berking C, Schilling B, Haferkamp S, Schadendorf D, Holland-Letz T, Utikal JS, von Kalle C; Collaborators. Deep learning outperformed 136 of 157 dermatologists in a head-to-head dermoscopic melanoma image classification task. Eur J Cancer. 2019 May;113:47–54. doi: 10.1016/j.ejca.2019.04.001. Epub 2019 Apr 10. PMID: 30981091.

Brinker, T. J., Hekler, A., Utikal, J. S., Grabe, N., Schadendorf, D., Klode, J., & Esser, S. (2018). Skin cancer classification using convolutional neural networks: systematic review. Journal of Medical Internet Research, 21(10), e11936.

Chicco, D., Jurman, G. The advantages of the Matthews correlation coefficient (MCC) over F1 score and accuracy in binary classification evaluation. BMC Genomics 21, 6 (2020). 10.1186/s12864-019-6413-7

Cristiano Patrício, João C. Neves, and Luís F. Teixeira. 2023. Explainable Deep Learning Methods in Medical Image Classification: A Survey. ACM Comput. Surv. 56, 4, Article 85 (April 2024), 41 pages. 10.1145/3625287

C. Pandey, A. D. Choudhury and S. Khandelwal, “qxAI: Quantifiable xAI for Cardiac Diseases,” 2024 IEEE International Conference on Pervasive Computing and Communications Workshops and other Affiliated Events (PerCom Workshops), Biarritz, France, 2024, pp. 233–238, doi: 10.1109/PerComWorkshops59983.2024.10502886.

Daneshjou R, Barata C, Betz-Stablein B, et al. Checklist for Evaluation of Image-Based Artificial Intelligence Reports in Dermatology: CLEAR Derm Consensus Guidelines From the International Skin Imaging Collaboration Artificial Intelligence Working Group. JAMA Dermatol. 2022;158(1):90–96. doi:10.1001/jamadermatol.2021.4915

Doshi-Velez, F., & Kim, B. (2017). Towards a rigorous science of interpretable machine learning. doi: 10.48550/arXiv.1702.08608

Ehsan, U., Harrison, B., Chan, L. and Riedl, M.O. (2017). Rationalization: A Neural Machine Translation Approach to Generating Natural Language Explanations. [online] arXiv.org. doi: 10.48550/arXiv.1702.07826.

Eloise Withnell, Xiaoyu Zhang, Kai Sun, Yike Guo, XOmiVAE: an interpretable deep learning model for cancer classification using high-dimensional omics data, Briefings in Bioinformatics, Volume 22, Issue 6, November 2021, bbab315, 10.1093/bib/bbab315

Esteva, A., Kuprel, B., Novoa, R. et al. Dermatologist-level classification of skin cancer with deep neural networks. Nature 542, 115–118 (2017). 10.1038/nature21056

European Commission. (2021). Proposal for a Regulation of the European Parliament and of the Council laying down harmonised rules on artificial intelligence (Artificial Intelligence Act) and amending certain Union legislative acts. COM/2021/206 final.

Fong, R. and Vedaldi, A. (2017). Interpretable Explanations of Black Boxes by Meaningful Perturbation. doi: 10.1109/iccv.2017.371

Ghanvatkar, S., and Rajan, V. “Evaluating Explanations From AI Algorithms for Clinical Decision-Making: A Social Science-Based Approach,” in IEEE Journal of Biomedical and Health Informatics, vol. 28, no. 7, pp. 4269–4280, July 2024, doi: 10.1109/JBHI.2024.3393719

Goodfellow, I., Bengio, Y., & Courville, A. (2016). Deep Learning. MIT Press.

Gloster, H.M. and Neal, K. (2006). Skin cancer in skin of color. Journal of the American Academy of Dermatology, [online] 55(5), pp.741–760. doi: 10.1016/j.jaad.2005.08.063.

Qi Han, Xin Qian, Hongxiang Xu, Kepeng Wu, Lun Meng, Zicheng Qiu, Tengfei Weng, Baoping Zhou, Xianqiang Gao, DM-CNN: Dynamic Multi-scale Convolutional Neural Network with uncertainty quantification for medical image classification, Computers in Biology and Medicine, Volume 168, 2024, 107758, ISSN 0010-4825, 10.1016/j.compbiomed.2023.107758.

Haenssle HA, Fink C, Schneiderbauer R, Toberer F, Buhl T, Blum A, Kalloo A, Hassen ABH, Thomas L, Enk A, Uhlmann L; Man against machine: diagnostic performance of a deep learning convolutional neural network for dermoscopic melanoma recognition in comparison to 58 dermatologists. Ann Oncol. 2018 Aug 1;29(8):1836–1842. doi: 10.1093/annonc/mdy166. PMID: 29846502.

He, K., Zhang, X., Ren, S., & Sun, J. (2016). Deep residual learning for image recognition. doi: 10.48550/arXiv.1512.03385

Hedström, A., Weber, L., Bareeva, D., Krakowczyk, D., Motzkus, F., Samek, W., Lapuschkin, S. and Höhne, C. (2023). Quantus: An Explainable AI Toolkit for Responsible Evaluation of Neural Network Explanations and Beyond. Journal of Machine Learning Research, [online] 24, pp.1–11. Available at: https://jmlr.org/papers/volume24/22-0142/22-0142.pdf [Accessed 13 Aug. 2024].

Hicks, S.A., Strümke, I., Thambawita, V. et al. On evaluation metrics for medical applications of artificial intelligence. Sci Rep 12, 5979 (2022). 10.1038/s41598-022-09954-8

Hossain MM, Hossain MM, Arefin MB, Akhtar F, Blake J. Combining State-of-the-Art Pre-Trained Deep Learning Models: A Noble Approach for Skin Cancer Detection Using Max Voting Ensemble. Diagnostics (Basel). 2023 Dec 30;14(1):89. doi: 10.3390/diagnostics14010089. PMID: 38201399; PMCID: PMC10795598.

Hu, B., B. Vasu and A. Hoogs, “X-MIR: EXplainable Medical Image Retrieval,” 2022 IEEE/CVF Winter Conference on Applications of Computer Vision (WACV), Waikoloa, HI, USA, 2022, pp. 1544–1554, doi: 10.1109/WACV51458.2022.00161.

Huang, G., Liu, Z., Van Der Maaten, L., & Weinberger, K. Q. (2017). Densely connected convolutional networks. doi: 10.48550/arXiv.1608.06993

Huang, H.-W., Hsu, B.W.-Y., Lee, C.-H. and Tseng, V.S. (2021), Development of a light-weight deep learning model for cloud applications and remote diagnosis of skin cancers. J. Dermatol., 48: 310–316. 10.1111/1346-8138.15683

Holzinger, A., Biemann, C., Pattichis, C. and Kell, D. (2017). What do we need to build explainable AI systems for the medical domain? [online] Available at: https://arxiv.org/pdf/1712.09923.pdf.

Howard, A. G., Zhu, M., Chen, B., Kalenichenko, D., Wang, W., Weyand, T., … & Adam, H. (2017). MobileNets: Efficient convolutional neural networks for mobile vision applications. doi: 10.48550/arXiv.1704.04861

International Skin Imaging Collaboration. SLICE-3D 2024 Challenge Dataset. International Skin Imaging Collaboration 10.34970/2024-slice-3d (2024).

International Organization for Standardization (ISO). (2016). ISO 13485:2016 Medical devices – Quality management systems – Requirements for regulatory purposes.

International Organization for Standardization (ISO). (2019). ISO 14971:2019 Medical devices – Application of risk management to medical devices.

International Organization for Standardization (ISO). (2015). ISO 14001:2015 Environmental management systems – Requirements with guidance for use.

International Organization for Standardization (ISO). (2008). ISO 9241-171:2008 Ergonomics of human-system interaction – Part 171: Guidance on software accessibility.

Jin, W., Li, X. and Hamarneh, G. (2022). Evaluating Explainable AI on a Multi-Modal Medical Imaging Task: Can Existing Algorithms Fulfill Clinical Requirements? Proceedings of the AAAI Conference on Artificial Intelligence, 36(11), pp.11945–11953. doi: 10.1609/aaai.v36i11.21452

Jin, W., Xiaoxiao Li, Mostafa Fatehi, Ghassan Hamarneh, Guidelines and evaluation of clinical explainable AI in medical image analysis, Medical Image Analysis, Volume 84, 2023, 102684, ISSN 1361-8415, 10.1016/j.media.2022.102684

M. Hosny, M. A. Kassem and M. M. Foaud, “Skin Cancer Classification using Deep Learning and Transfer Learning,” 2018 9th Cairo International Biomedical Engineering Conference (CIBEC), Cairo, Egypt, 2018, pp. 90–93, doi: 10.1109/CIBEC.2018.8641762.

Kumar Lilhore, U., Simaiya, S., Sharma, Y.K. et al. A precise model for skin cancer diagnosis using hybrid U-Net and improved MobileNet-V3 with hyperparameters optimization. Sci Rep 14, 4299 (2024). 10.1038/s41598-024-54212-8

Kingma, D. P., & Ba, J. (2014). Adam: A Method for Stochastic Optimization. doi: 10.48550/arXiv.1412.6980

L. N. Smith, “Cyclical Learning Rates for Training Neural Networks,” 2017 IEEE Winter Conference on Applications of Computer Vision (WACV), Santa Rosa, CA, USA, 2017, pp. 464–472, doi: 10.1109/WACV.2017.58.

Ladbury, C., Reza Zarinshenas, Hemal Semwal, Tam, A., Nagarajan Vaidehi, Rodin, A.S., Liu, A., Glaser, S., Ravi Salgia and Amini, A. (2022). Utilization of model-agnostic explainable artificial intelligence frameworks in oncology: a narrative review. Transatlantic Cancer Research, 11(10), pp.3853–3868. doi: 10.21037/tcr-22-1626.

Li, Y., Zhou, J., Verma, S. and Chen, F. (2023). A Survey of Explainable Graph Neural Networks: Taxonomy and Evaluation Metrics. [online] Available at: https://arxiv.org/pdf/2207.12599.pdf [Accessed 9 Feb. 2024].

Luis A. de Souza, Robert Mendel, Sophia Strasser, Alanna Ebigbo, Andreas Probst, Helmut Messmann, João P. Papa, Christoph Palm, Convolutional Neural Networks for the evaluation of cancer in Barrett’s esophagus: Explainable AI to lighten up the black-box, Computers in Biology and Medicine, Volume 135, 2021, 104578, ISSN 0010-4825, 10.1016/j.compbiomed.2021.104578

Lundberg, S. M., & Lee, S. I. (2017). A unified approach to interpreting model predictions. doi: 10.48550/arXiv.1705.07874

Md. Kamrul Hasan, Md. Toufick E. Elahi, Md. Ashraful Alam, Md. Tasnim Jawad, Robert Martí, DermoExpert: Skin lesion classification using a hybrid convolutional neural network through segmentation, transfer learning, and augmentation, Informatics in Medicine Unlocked, Volume 28, 2022, 100819, ISSN 2352-9148, 10.1016/j.imu.2021.100819.

Md. Aminur Rab Ratul, M. Hamed Mozaffari, Won-Sook Lee, Enea Parimbelli, Skin Lesions Classification Using Deep Learning Based on Dilated Convolution, bioRxiv 860700; doi: 10.1101/860700

M. Sobhan and A. M. Mondal, “Explainable Machine Learning to Identify Patient-specific Biomarkers for Lung Cancer,” 2022 IEEE International Conference on Bioinformatics and Biomedicine (BIBM), Las Vegas, NV, USA, 2022, pp. 3152–3159, doi: 10.1109/BIBM55620.2022.9995516.

Magesh, P.R., Myloth, R.D. and Tom, R.J. (2020). An Explainable Machine Learning Model for Early Detection of Parkinson’s Disease using LIME on DaTSCAN Imagery. Computers in Biology and Medicine, 126, p.104041. doi: 10.1016/j.compbiomed.2020.104041

Makridis et al., “Towards a Unified Multidimensional Explainability Metric: Evaluating Trustworthiness in AI Models,” 2023 19th International Conference on Distributed Computing in Smart Systems and the Internet of Things (DCOSS-IoT), Pafos, Cyprus, 2023, pp. 504–511, doi: 10.1109/DCOSS-IoT58021.2023.00084.

Miller, T., Explanation in artificial intelligence: Insights from the social sciences, Artificial Intelligence, Volume 267, 2019, Pages 1–38, ISSN 0004-3702, 10.1016/j.artint.2018.07.007.

Mohan, H. and Yoo, J. (2023). A comprehensive analysis of recent advancements in cancer detection using machine learning and deep learning models for improved diagnostics. Journal of Cancer Research and Clinical Oncology, 149(15), pp.14365–14408. doi:10.1007/s00432-023-05216-w.

Mukherjee, S., Adhikari, A. and Roy, M. (2019). Malignant Melanoma Classification Using Cross-Platform Dataset with Deep Learning CNN Architecture. Recent Trends in Signal and Image Processing, pp.31–41. doi:10.1007/978-981-13-6783-0_4.

Nitesh V. Chawla, Kevin W. Bowyer, Lawrence O. Hall, and W. Philip Kegelmeyer. 2002. SMOTE: synthetic minority over-sampling technique. J. Artif. Int. Res. 16, 1 (January 2002), 321–357.

Pieter Van Molle, Miguel De Strooper, Verbelen, T., Vankeirsbilck, B., Simoens, P. and Dhoedt, B. (2018). Visualizing Convolutional Neural Networks to Improve Decision Support for Skin Lesion Classification. Lecture Notes in Computer Science, pp.115–123. doi:10.1007/978-3-030-02628-8_13.

Perez, L., & Wang, J. (2017). The effectiveness of data augmentation in image classification using deep learning. arXiv preprint 1712.04621.

Poursabzi-Sangdeh, F., Goldstein, D.G., Hofman, J.M., Vaughan, J.W. and Wallach, H. (2021). Manipulating and Measuring Model Interpretability. 1802.07810 [cs]. [online] Available at: https://arxiv.org/abs/1802.07810.

Priti Bansal, Ritik Garg, Priyank Soni, Detection of melanoma in dermoscopic images by integrating features extracted using handcrafted and deep learning models, Computers & Industrial Engineering, Volume 168, 2022, 108060, ISSN 0360-8352, 10.1016/j.cie.2022.108060.

Rajendran, B., Simeone, O. and Al-Hashimi, B. (2023). Towards Efficient and Trustworthy AI Through Hardware-Algorithm-Communication Co-Design. [online] Available at: https://arxiv.org/pdf/2309.15942.pdf [Accessed 8 Feb. 2024].

Rezazadeh, A., Jafarian, Y. and Kord, A. (2022). Explainable Ensemble Machine Learning for Breast Cancer Diagnosis Based on Ultrasound Image Texture Features. Forecasting, 4(1), pp.262–274. doi: 10.3390/forecast4010015.

Ribeiro, M. T., Singh, S., & Guestrin, C. (2016). “Why should I trust you?” Explaining the predictions of any classifier. doi: 10.48550/arXiv.1602.04938

Rudin, C. Stop explaining black box machine learning models for high stakes decisions and use interpretable models instead. Nat Mach Intell 1, 206–215 (2019). 10.1038/s42256-019-0048-x

Salih O, Duffy KJ. Optimization Convolutional Neural Network for Automatic Skin Lesion Diagnosis Using a Genetic Algorithm. Applied Sciences. 2023; 13(5):3248. 10.3390/app13053248

Sangwan H., 2024, Github Repository, https://github.com/hardikSangwan/thesis_diagnostics_skin

Selvaraju, R. R., Cogswell, M., Das, A., Vedantam, R., Parikh, D., & Batra, D. (2017). Grad-CAM: Visual explanations from deep networks via gradient-based localization. doi: 10.48550/arXiv.1610.02391

Shahsavari, A., Khatibi, T. & Ranjbari, S. Skin lesion detection using an ensemble of deep models: SLDED. Multimed Tools Appl 82, 10575–10594 (2023). 10.1007/s11042-022-13666-6

Siddiqui, K. and T. E. Doyle, “Trust Metrics for Medical Deep Learning Using Explainable-AI Ensemble for Time Series Classification,” 2022 IEEE Canadian Conference on Electrical and Computer Engineering (CCECE), Halifax, NS, Canada, 2022, pp. 370–377, doi: 10.1109/CCECE49351.2022.9918458.

Singh, A.; Sengupta, S.; Lakshminarayanan, V. Explainable Deep Learning Models in Medical Image Analysis. J. Imaging 2020, 6, 52. 10.3390/jimaging6060052

Sundararajan, M., Taly, A., & Yan, Q. (2017). Axiomatic attribution for deep networks, https://arxiv.org/abs/1703.01365

Szegedy, C., Ioffe, S., Vanhoucke, V., & Alemi, A. (2017). Inception-v4, inception-resnet and the impact of residual connections on learning. doi: 10.48550/arXiv.1602.07261

T. Alkarakatly, S. Eidhah, M. Al-Sarawani, A. Al-Sobhi and M. Bilal, “Skin Lesions Identification Using Deep Convolutional Neural Network,” 2019 International Conference on Advances in the Emerging Computing Technologies (AECT), Al

Tabrizchi, H., Parvizpour, S. & Razmara, J. An Improved VGG Model for Skin Cancer Detection. Neural Process Lett 55, 3715–3732 (2023). 10.1007/s11063-022-10927-1

Todd Kulesza, Margaret Burnett, Weng-Keen Wong, and Simone Stumpf. 2015. Principles of Explanatory Debugging to Personalize Interactive Machine Learning. In Proceedings of the 20th International Conference on Intelligent User Interfaces (IUI ‘15). Association for Computing Machinery, New York, NY, USA, 126–137. 10.1145/2678025.2701399

Tschandl, Philipp, 2018, “The HAM10000 dataset, a large collection of multi-source dermatoscopic images of common pigmented skin lesions”, 10.7910/DVN/DBW86T, Harvard Dataverse, V4

United Nations Sustainable Development. Home - 2019 - United Nations Sustainable Development. [online] Available at: https://www.un.org/sustainabledevelopment.

U.S. Food and Drug Administration (FDA). (2021). Artificial Intelligence and Machine Learning (AI/ML) Software as a Medical Device Action Plan.

Wang, C. (2023). Calibration in Deep Learning: A Survey of the State-of-the-Art. [online] Available at: https://arxiv.org/pdf/2308.01222.pdf [Accessed 8 Feb. 2024].

Xu, Y., Lam, H.-K. and Jia, G. (2021). MANet: A two-stage deep learning method for classification of COVID-19 from Chest X-ray images. Neurocomputing, 443, pp.96–105. doi: 10.1016/j.neucom.2021.03.034

Y. Bengio, Neural Networks: Tricks of the Trade chapter Practical recommendations for gradient-based training of deep architectures, Springer Berlin Heidelberg, pp. 437–478, 2012.

Y. Jiang, S. Cao, S. Tao and H. Zhang, “Skin Lesion Segmentation Based on Multi-Scale Attention Convolutional Neural Network,” in IEEE Access, vol. 8, pp. 122811–122825, 2020, doi: 10.1109/ACCESS.2020.3007512.

Yaqoob, A., Rabia Musheer Aziz and Navneet Kumar Verma (2023). Applications and Techniques of Machine Learning in Cancer Classification: A Systematic Review. Human-Centric Intelligent Systems. doi: 10.1007/s44230-023-00041-3.

Yosinski, J., Jeff Clune, Yoshua Bengio, and Hod Lipson. 2014. How transferable are features in deep neural networks? In Proceedings of the 27th International Conference on Neural Information Processing Systems - Volume 2 (NIPS’14). MIT Press, Cambridge, MA, USA, 3320–3328.

Zhang, Li, Jian Zhang, Wenlian Gao, Fengfeng Bai, Nan Li, Noradin Ghadimi, A deep learning outline aimed at prompt skin cancer detection utilizing gated recurrent unit networks and improved orca predation algorithm, Biomedical Signal Processing and Control, Volume 90, 2024, 105858, ISSN 1746-8094, 10.1016/j.bspc.2023.105858.

Zhiwei Qin, Zhao Liu, Ping Zhu, Yongbo Xue, A GAN-based image synthesis method for skin lesion classification, Computer Methods and Programs in Biomedicine, Volume 195, 2020, 105568, ISSN 0169-2607, 10.1016/j.cmpb.2020.105568.

Zou, L. et al., “Ensemble Image Explainable AI (XAI) Algorithm for Severe Community-Acquired Pneumonia and COVID-19 Respiratory Infections,” in IEEE Transactions on Artificial Intelligence, vol. 4, no. 2, pp. 242–254, April 2023, doi: 10.1109/TAI.2022.3153754

